# Adiposity and Mortality among Patients Severely Ill with COVID-19 and non-COVID-19 Respiratory Conditions: A Cross-Context Comparison Study in the UK

**DOI:** 10.1101/2022.12.22.22283842

**Authors:** Joshua A. Bell, David Carslake, Amanda Hughes, Kate Tilling, James W. Dodd, James C. Doidge, David A. Harrison, Kathryn M. Rowan, George Davey Smith

## Abstract

**Objective:** To assess the causality of adiposity for mortality among patients severely ill with COVID-19 and non-COVID-19 respiratory conditions by examining the consistency of associations across temporal and geographical contexts where biases vary

**Design:** Prospective cohort study

**Setting:** 297 intensive care units (ICUs) in England, Wales, and Northern Ireland monitored by the Intensive Care National Audit and Research Centre Case Mix Programme

**Participants:** Patients aged ≥16 years admitted to ICU with COVID-19 (N=33,352; Feb 2020-Aug 2021) and non-COVID-19 respiratory conditions (N=24,739; Feb 2018-Aug 2019)

**Main outcome measure:** 30-day mortality post ICU admission

**Results:** Compared with non-COVID-19 respiratory patients, COVID-19 patients were younger, less often of a white ethnic group, and more often with extreme obesity (body mass index (BMI) ≥ 40kg/m^2^). COVID-19 patients had fewer comorbidities but higher mortality (35% vs. 23% mortality in non-COVID-19). Socio-demographic and comorbidity factors and their associations with BMI and mortality varied more by date than geographical region of ICU admission, particularly among COVID-19 patients. Among COVID-19 patients, higher BMI was associated with a small excess mortality (hazard ratio (HR) per standard deviation (SD)=1.05; 95% CI=1.03, 1.08), driven by extreme obesity (HR per SD=1.21; 95% CI=1.13, 1.31 vs. normal-weight). Extreme obesity was strongly associated with higher mortality only during Feb-April 2020 (HR=1.49, 95% CI=1.27, 1.73 vs. normal-weight); this association weakened thereafter (BMI-date interaction P=0.03). Among non-COVID-19 respiratory patients, higher BMI was associated with lower mortality (HR per SD=0.84; 95% CI=0.82, 0.87), seen across all overweight/obesity groups. These negative obesity-mortality associations were similar across most admission dates and regions.

**Conclusions:** Obesity is associated with higher mortality among COVID-19 patients, but lower mortality among non-COVID respiratory patients. These associations appear vulnerable to confounding/selection bias in both patient groups, questioning the existence or stability of causal effects. Among COVID-19 patients, unfavourable obesity-mortality associations differ by admission date. Among non-COVID-19 respiratory patients, favourable obesity-mortality associations may reflect comorbidity-induced weight loss.

## Background

The global spread of severe acute respiratory syndrome coronavirus 2 (SARS-CoV-2), and the resultant coronavirus disease 2019 (COVID-19), continues to threaten public health (1). Identifying modifiable causes of mortality among patients severely ill with COVID-19 remains a priority. In the post-2022 era, this task coincides with the need to manage possible dual surges of severe COVID-19 and influenza-related respiratory diseases, and studies must now consider the impact of risk factors within both conditions to guide appropriate messaging. Higher adiposity likely causes numerous non-infectious diseases (2) and large-scale evidence now supports adiposity as a potential cause of unfavourable COVID-19-related outcomes including initial SARS-CoV-2 infection (3), hospitalisation with severe COVID-19, and higher mortality with COVID-19 (3-9). This contrasts sharply with evidence on patients with non-COVID-19 respiratory conditions, with prior studies suggesting that higher adiposity including extreme obesity carries *lower* mortality (10, 11). However, potential effects of adiposity among patients with severe respiratory disease have mostly been estimated via conventional observational studies which use multivariable adjustments to address confounding and other biases. Mendelian randomization has been applied to population-based samples but is often unfeasible in hospital settings. As a result, the potential for unmeasured/residual confounding, reverse causation, and selection bias makes causality difficult to infer.

Cross-context comparison is an underutilised tool for causal inference in observational studies. This approach involves directly comparing associations between exposures and outcomes across temporal or geographical contexts where confounding or selection pressures vary. Consistency in the direction and magnitude of exposure-outcome associations across contexts, despite variation in the impact of confounding/selection, builds confidence in the causality of those exposure-outcome associations (12, 13). Previously, comparisons across geographical contexts where socioeconomic gradients in exposures differ helped to affirm the likely effects of gestational blood glucose on offspring birthweight and of breastfeeding on offspring intelligence, and reduce confidence in the suggested benefits of breastfeeding for offspring adiposity and blood pressure (12, 14). Cross-context comparisons have not been formally applied to assess the causality of adiposity for mortality among patients severely ill with COVID-19 and non-COVID-19 respiratory conditions, but this is now feasible within the UK setting given that the prevalence and impact of confounding/selection factors among patients may differ by time and geography (15-17).

Cross-context comparison requires variation in bias across contexts (time/place), i.e., it can assess the impact of context-varying bias, but not the impact of context-stable bias. Given uniquely rapid changes in COVID-19 management over time and geography, we may expect context-varying bias to influence adiposity-mortality associations more among COVID-19 patients than non-COVID-19 respiratory patients. Moreover, the influence of context-stable bias, e.g., reverse causation, may influence adiposity-mortality associations more among non-COVID-19 respiratory patients. This may be assessable by comparing characteristics of COVID-19 and non-COVID-19 respiratory patients and examining how adiposity-mortality associations differ between them.

The Intensive Care National Audit and Research Centre (ICNARC) has coordinated the collection of data on nearly all patients admitted to intensive care units (ICUs) in England, Wales, and Northern Ireland since 2010 (15, 16, 18, 19). In this study, we aimed to assess the causality of adiposity for mortality among people hospitalised with severe COVID-19 and non-COVID-19 respiratory conditions using a cross-context comparison approach. We used nationally representative data from the ICNARC Case Mix Programme on patients admitted to ICU with COVID-19 (∼33,000 patients between Feb 2020-Aug 2021) and with non-COVID-19 respiratory conditions (∼25,000 patients between Feb 2018-Aug 2019 (pre-pandemic)). Within each patient group, we estimated the overall association between adiposity and mortality. We then examined whether socio-demographic and comorbidity indicators (potential confounding/selection factors) and their associations with adiposity and mortality vary by date and geographical region of ICU admission. Lastly, we examined whether adiposity-mortality associations are consistent across dates and regions with varying confounding/selection pressures.

## Methods

This cohort study is reported in accordance with the Strengthening the Reporting of Observational Studies in Epidemiology (STROBE) guidelines (checklist in **Supp Item 1**).

### Study population

We included patients aged ≥ 16 years admitted to any of 280 ICUs across England, Wales, and Northern Ireland with COVID-19 confirmed at or after admission between 5 Feb 2020 and 1 Aug 2021, plus adult patients admitted to any of 266 ICUs with respiratory diseases which were not COVID-19, including viral and bacterial pneumonia, bronchitis, bronchiolitis, or laryngotracheobronchitis (encompassing suspected/confirmed influenza) between 1 Feb 2018 and 31 Aug 2019 (**Figure 1**). These start/end dates for COVID-19 admissions were based on data availability when this study commenced; the start/end dates for non-COVID-19 admissions were chosen to match the first/last months of COVID-19 admissions in a similarly long pre-pandemic period. Non-overlapping periods were used for primary analyses because these were expected to involve less disease misclassification and less estimate imprecision for non-COVID-19 respiratory conditions given their relative rarity during COVID-19 waves (e.g., given lockdowns). Data were from ICUs participating in the Case Mix Programme: the national clinical audit covering all National Health Service (NHS) adult, general intensive care, and combined intensive care/high dependency units, plus some additional specialist ICUs and standalone high dependency units, coordinated by ICNARC (18-20). This audit excludes all paediatric and neonatal ICUs and ICUs in Scotland. In cases where more than one ICU admission record exists, data from the most recent admission and discharge were used. Approval for the collection and use of patient-identifiable data without consent in the Case Mix Programme was obtained from the Confidentiality Advisory Group of the Health Research Authority under Section 251 of the NHS Act 2006 (approval number PIAG2–10[f]/2005). All data were pseudonymised (patient identifiers removed) prior to extraction for this research.

**Figure 1.**
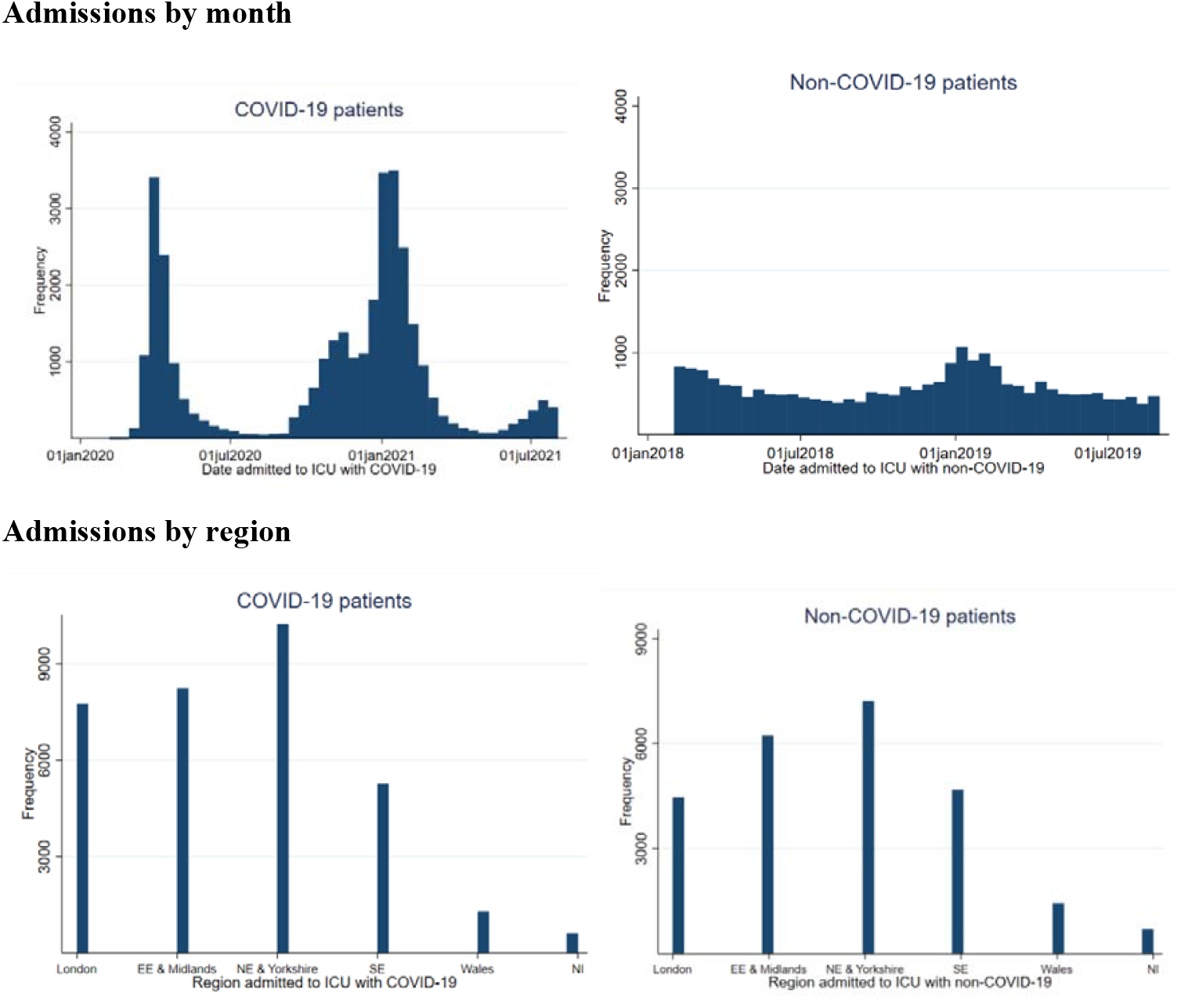
Numbers of patients admitted to any of 280 ICUs in England, Wales, and Northern Ireland with COVID-19 (5 Feb 2020 to 1 Aug 2021) and to any of 266 ICUs with non-COVID-19 respiratory conditions (1 Feb 2018 to 31 Aug 2019) participating in the ICNARC Case Mix Programme (297 ICUs overall) **Note:** Numbers are restricted to those with data on BMI, ≥1 sociodemographic/comorbidity indicator, and mortality and thus who are eligible for ≥1 analysis. EE = East England. NE = North-East England. SE = South-East England. NI = Northern Ireland.

### Adiposity (exposure)

Body mass index (BMI, as kg/m^2^) was calculated from height and weight which were recorded or estimated by clinicians on admission to ICU. Patient severity and clinician workload often make directly measuring height and weight in ICU infeasible (21). About half of the height and weight recordings were estimated rather than directly measured for both COVID-19 and non-COVID-19 respiratory patients (54% of each had at least one of height and weight estimated, and 38% of COVID-19 patients and 37% of non-COVID-19 respiratory patients had both height and weight estimated). Previous analyses of ICNARC ICU data supported similar associations of BMI with mortality based on measured vs. estimated values of height and weight (22), and we therefore expected little impact of this measurement error in BMI on BMI-mortality associations aside from potential bias towards the null. We examined BMI as a continuous variable (per standard deviation (SD) based on z-scores derived separately within COVID-19 and non-COVID-19 groups) and as a categorical variable based on World Health Organization classifications of underweight (<18.5 kg/m^2^), normal-weight (18.5-24.9 kg/m^2^), overweight (25.0-29.9 kg/m^2^), obesity class 1 (30.0-34.9 kg/m^2^), obesity class 2 (35.0-39.9 kg/m^2^), and obesity class 3+ (≥40.0 kg/m^2^).

### 30-day all-cause mortality (outcome)

We included deaths in ICU from any cause (reported directly to ICNARC by hospital staff). We censored deaths at 30 days post-admission to ICU and assumed that patients discharged from ICU lived to be censored at 30 days post-admission (15). This was because censoring discharged patients at discharge would have selectively removed the healthiest patients from follow-up (‘informative’ censoring). Deaths occurring more than 30 days post-admission were excluded due to the generally high short-term mortality rate within ICU settings; later deaths are rare and risk introducing unrelated causes of death, while the assumption of survival in discharged patients also becomes less plausible with time (23).

### Socio-demographic and comorbidity indicators (confounding and selection factors)

Measured confounders (hypothesised to cause adiposity and mortality) included patient age, sex, ethnic group, socioeconomic deprivation, and the geographical region and date of ICU admission. Age was used as a continuous variable (assumed to have a linear effect on log-survival). Ethnic groups included ‘White’, ‘Black’, ‘Asian’ (specifically South Asian), ‘mixed/other’ (including East Asian), based on 2011 census categories used for NHS data entry. Deprivation was based on quintiles of the Index of Multiple Deprivation (IMD) which summarises several area-level deprivation indicators including income, education, and employment for neighbourhood-level areas within each UK country, derived from patients’ postcodes. Geographic region was grouped as: 1) London; 2) East England and Midlands; 3) North-East England, North-West England, and Yorkshire; 4) South-East England and South-West England, 5) Wales; and 6) Northern Ireland. Admission date was considered as 6 groups for COVID-19 patients: 1) 5 Feb-31 April 2020; 2) 1 May-31 July 2020; 3) 1 Aug-31 Oct 2020; 4) 1 Nov-31 Jan 2021; 5) 1 Feb-31 April 2021; and 6) 1 May-1 Aug 2021; and 6 groups for non-COVID-19 respiratory patients: 1) 1 Feb-31 April 2018; 2) 1 May-31 July 2018; 3) 1 Aug-31 Oct 2018; 4) 1 Nov-31 Jan 2019; 5) 1 Feb-31 April 2019; and 6) 1 May-31 Aug 2019.

Several comorbidity indicators were also recorded and considered here primarily as selection factors (not confounders for multivariable adjustment). These included severe comorbid diseases documented in the patient notes as being present within the 6 months prior to ICU admission: respiratory disease (shortness of breath with light activity or home ventilation), cardiovascular disease (symptoms at rest), end-stage renal disease requiring chronic renal replacement therapy, liver disease (biopsy-proven cirrhosis, portal hypertension or hepatic encephalopathy), metastatic disease, haematological disease (acute or chronic leukaemia, multiple myeloma, or lymphoma) and an immunocompromised state (chemotherapy, radiotherapy, daily high-dose steroids, AIDS, or acquired immunohumoral or cellular immune deficiency). As indicators of acute severity within ICU, we included: 1) the acute physiology and chronic health evaluation-II (APACHE-II) score and the ICNARC extreme physiology score which summarise patient physiology within the first 24 hours in ICU (higher values being adverse), 2) the ratio of arterial oxygen partial pressure (PaO2) to fractional inspired oxygen (FiO2) (kPa units) calculated from arterial blood gas with the lowest PaO2 in the first 24 hours, and 3) the number of days on which advanced respiratory support was received (including ventilation). Additionally, we examined two indicators of pre-ICU health status: whether the patient was physically dependent on others for activities of daily living prior to admission (some dependency; total dependency; or independent), and whether the patient had any past severe illness (defined as having a zero value of the APACHE-II past medical history weighting, grouped as yes/no).

### Statistical approach

#### 1. Analyses informing cross-context comparisons

We described the socio-demographic and comorbidity indicators (confounding/selection factors) of patients admitted to ICU with COVID-19 and non-COVID-19 respiratory conditions via proportions or means and SDs. Patient groups were first described overall, and then with separate stratification by month and region of ICU admission to examine temporal/geographic trends in patient characteristics. These temporal trend descriptions excluded COVID-19 patients admitted during Feb 2020 and Aug 2021 due to low case counts (5 and 3 cases respectively); these patients were not excluded from subsequent regression models because those models use aggregated cases/months only.

Separately among COVID-19 and non-COVID-19 respiratory patients, we used regression models to examine associations of socio-demographic and comorbidity indicators (independent variables in separate models) with BMI using linear models, and with mortality using Weibull proportional hazards models, each adjusted for age and sex. Weibull models were used as a parametric alternative to semi-parametric Cox models to relax the unrealistic assumption of equivalent baseline hazards when later stratifying models on patient group and ICU admission month and region. Time since ICU admission was used as the time axis. We repeated these separate models with stratification on month and region of ICU admission, each in 6 groups as described above. Evidence of statistical heterogeneity was tested between patient group (COVID-19 vs. non-COVID-19) and each confounding/selection factor, and likewise for date (6 groups) and region (6 groups) and with each confounding/selection factor (using the post-estimation ‘test’ command in Stata) in relation to BMI and mortality as outcomes using separate linear or Weibull models, respectively.

#### 2. Analyses for overall adiposity-mortality associations and cross-context comparisons

We used Weibull regression models to estimate the overall association of BMI, in SD units and categorically relative to normal-weight, with 30-day all-cause mortality among ICU patients with COVID-19 and non-COVID-19 respiratory conditions, adjusted for confounders of age, sex, ethnic group, deprivation, admission date, and admission region. The time axis, entry, and censoring for these Weibull models were as described above. The proportional hazards assumption was tested for these main Weibull models by splitting follow-up time based on the number of days marking the median time to death (4 days for COVID-19 patients and 10 days for non-COVID-19 patients) and comparing hazard ratios (HRs) between these 2 periods. These mortality HRs were similar between follow-up periods for each patient group, suggesting no strong evidence against proportionality. As sensitivity analyses for non-COVID-19 respiratory patients we: 1) additionally excluded patients admitted to ICU with bacterial pneumonia (thus considering only viral respiratory conditions for comparison with COVID-19), and 2) considered non-COVID-19 respiratory patients who were admitted during the same months as COVID-19 patients (1 Feb 2020 to 31 Aug 2021).

We then repeated Weibull models of BMI with mortality among COVID-19 and non-COVID-19 respiratory patients with the same confounding adjustments but with stratification by (instead of adjustment for) admission date and region using the groupings noted above. We tested interaction between BMI and admission date group, and between BMI and admission region group, in relation to mortality (smaller P-values indicate stronger evidence that associations vary by date/region), using the same post-estimation comparisons noted above. Analyses were conducted using Stata 16.

## Results

### Overall characteristics of ICU patients with COVID-19 and non-COVID-19 respiratory conditions

Of 39,431 adult patients admitted to ICU with COVID-19 between 5 Feb 2020 and 1 Aug 2021, 33,352 (85%) were eligible for ≥ 1 analysis based on having data on BMI (exposure), ≥ 1 sociodemographic/comorbidity indicator (confounding/selection factor), and 30-day ICU mortality (outcome) (**Figure 2**). Of 27,162 adult patients admitted to ICU with non-COVID-19 respiratory conditions between 1 Feb 2018 to 31 Aug 2019, 24,739 (91%) were eligible for ≥ 1 analysis based on the same criteria. For both patient groups, data were most often missing for confounding/selection factors.

**Figure 2.**
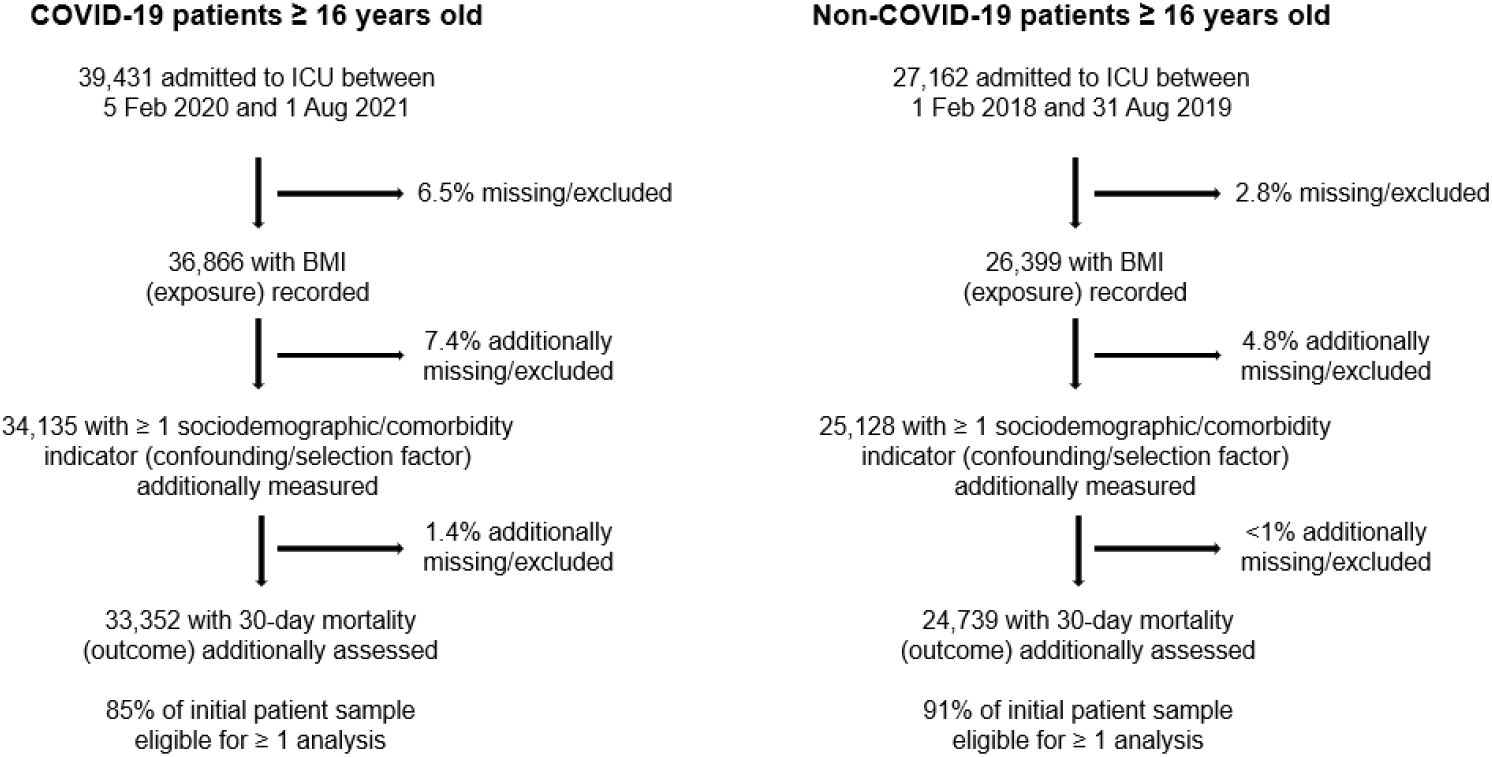
Flow of patients admitted to ICU with COVID-19 and non-COVID-19 respiratory conditions who are eligible for ≥ 1 analysis

Compared with non-COVID-19 patients, COVID-19 patients were younger, more often male, and substantially less often of a white ethnic group, with a more than two-fold higher proportion of Black, and more than three-fold higher proportion of Asian and mixed, ethnic groups (**Table 1**). COVID-19 patients were more often of the most deprived fifth of the IMD. COVID-19 patients had a higher mean BMI, at 30.9 kg/m^2^ vs. 27.6 kg/m^2^ in non-COVID-19 patients, and COVID-19 patients presented as underweight one-fifth as often and as normal-weight nearly half as often, yet presented with obesity, particularly classes 2 and 3+ obesity, about twice as often. COVID-19 patients had less prior severe illness than non-COVID-19 patients (7.8% vs. 21.3%) and less pre-ICU dependency on others for activities of daily living (11.3% vs. 33.9%). COVID-19 patients also presented less often with severe comorbidities including cardiovascular disease and liver, metastatic, haematological, respiratory, and immunocompromising diseases than non-COVID-19 patients. COVID-19 patients had a lower PaO2/FiO2 ratio, indicating worse respiratory function, but lower overall severity of illness as indicated by mean APACHE-II and ICNARC scores. Despite being younger with fewer comorbidities, COVID-19 patients experienced higher 30-day ICU mortality at 34.8% vs. 22.5% for non-COVID-19 patients.

**Table 1.**
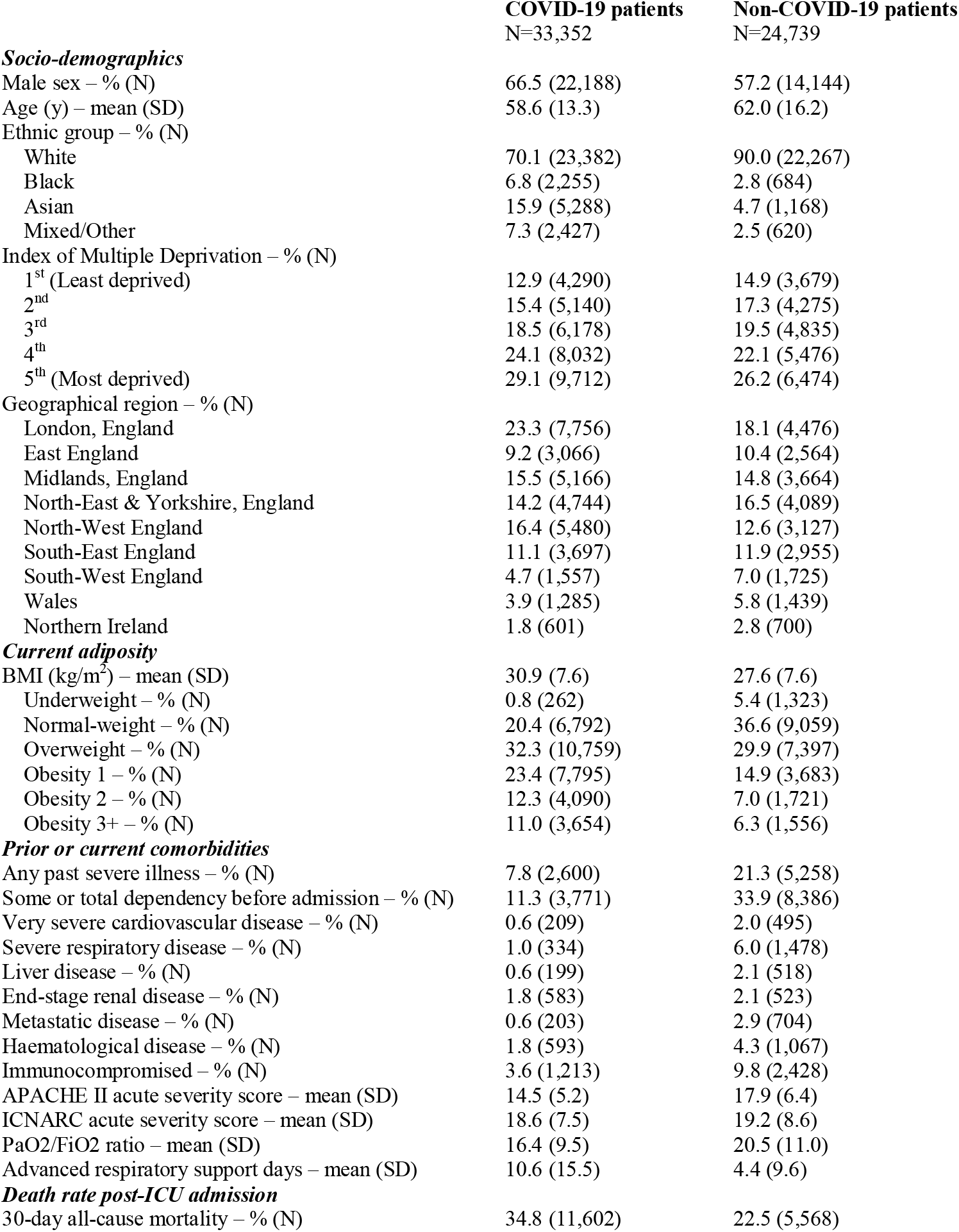
Characteristics of ICU patients in England, Wales, and Northern Ireland with COVID-19 (5 Feb 2020 to 1 Aug 2021) and non-COVID-19 respiratory conditions (1 Feb 2018 to 31 Aug 2019) eligible for analyses

### Overall associations of confounding/selection factors with BMI and mortality among ICU patients with COVID-19 and non-COVID-19 respiratory conditions

Among COVID-19 patients, each non-white vs. white ethnic group was associated with lower BMI, particularly for Asian ethnic group at -3.06 (95% CI=-3.27, -2.85) kg/m^2^ (**Supp Table 1**). These associations were weaker in non-COVID-19 patients. Higher deprivation was similarly associated with higher BMI in COVID-19 and non-COVID-19 patients. Most comorbidities were associated with lower BMI in both patient groups, but magnitudes were often higher among COVID-19 patients. In contrast, pre-ICU dependency and comorbid respiratory disease were associated with higher BMI in both groups.

Regarding mortality, associations of Black vs. white ethnic group were directionally opposite within patient groups (HR=1.10, 95% CI=1.02, 1.18 among COVID-19 patients vs. HR=0.67, 95% CI=0.55, 0.81 among non-COVID-19 patients). Asian vs. white ethnic group was associated with higher mortality among COVID-19 patients but was not associated with mortality among non-COVID-19 patients, whereas mixed vs. white ethnic group was associated with lower mortality in both patient groups. Deprivation was positively associated with mortality among COVID-19 patients only. Comorbidities were generally consistently associated with higher mortality across patient groups, except for end-stage renal disease which was associated with higher mortality among COVID-19 patients (HR=1.37, 95% CI=1.22, 1.55) yet lower mortality among non-COVID-19 patients (HR=0.80, 95% CI=0.66, 0.98).

### Overall associations of BMI with mortality among ICU patients with COVID-19 and non-COVID-19 respiratory conditions

Among COVID-19 patients admitted to ICU, higher BMI (per SD, or 7.6 kg/m^2^) was associated with a small excess ICU mortality (HR=1.05; 95% CI=1.03, 1.08; **Table 2**). Compared with normal-weight patients, underweight patients carried an excess mortality at HR=1.21 (95% CI=0.99, 1.48), whereas there was weak evidence of association of overweight, class 1 obesity, or 2 obesity with ICU mortality. Mortality was elevated with class 3+ obesity vs. normal-weight at HR=1.21 (95% CI=1.13, 1.31).

**Table 2.** Overall associations of BMI with 30-day all-cause mortality among ICU patients with COVID-19 (5 Feb 2020 to 1 Aug 2021) and non-COVID-19 respiratory conditions (1 Feb 2018 to 31 Aug 2019)

|  | **HR (95% CI) for 30-day all-cause mortality** |  |  |  |
| --- | --- | --- | --- | --- |
|  | **COVID-19 patients**<br>N=33,810 | **Non-COVID-19 patients**<br>N=24,879 | **Non-COVID-19 patients**<br>**excluding bacterial**<br>**pneumonia\***<br>N=19,275 | **Non-COVID-19 patients**<br>**admitted Feb 2020 to**<br>**Aug 2021\***<br>N=8,241 |
| **Adiposity (BMI)** |  |  |  |  |
| *Per SD higher BMI* | *1.05 (1.03, 1.08)* | *0.84 (0.82, 0.87)* | *0.84 (0.82, 0.87)* | *0.91 (0.87, 0.95)* |
| Underweight | 1.21 (0.99, 1.48) | 1.52 (1.36, 1.69) | 1.52 (1.35, 1.72) | 1.41 (1.15, 1.72) |
| Normal-weight | 1.00 (reference) | 1.00 (reference) | 1.00 (reference) | 1.00 (reference) |
| Overweight | 1.00 (0.96, 1.05) | 0.84 (0.79, 0.90) | 0.85 (0.79, 0.91) | 0.88 (0.78, 0.98) |
| Obesity 1 | 0.96 (0.91, 1.02) | 0.73 (0.67, 0.80) | 0.76 (0.69, 0.83) | 0.78 (0.67, 0.90) |
| Obesity 2 | 0.97 (0.91, 1.04) | 0.73 (0.64, 0.82) | 0.75 (0.65, 0.85) | 0.76 (0.63, 0.93) |
| Obesity 3+ | 1.21 (1.13, 1.31) | 0.73 (0.64, 0.83) | 0.70 (0.61, 0.82) | 0.92 (0.75, 1.12) |

Models are adjusted for sex, age, ethnic group, deprivation, geographical region, and admission date. \*Sensitivity analyses for non-COVID-19

Among non-COVID-19 patients admitted to ICU, higher BMI (per SD, or 7.3 kg/m^2^) was associated with lower ICU mortality (HR=0.84; 95% CI=0.82, 0.87). Compared with normal-weight patients, those who were underweight had substantial excess mortality at HR=1.52 (95% CI=1.36, 1.69), whereas patients who were overweight or with obesity had substantially lower mortality (HR=0.73; 95% CI=0.64, 0.83 for class 3+ obesity).

The pattern and magnitude of these associations were similar when excluding non-COVID-19 patients admitted to ICU with bacterial pneumonia (thus considering only viral respiratory conditions) and when considering non-COVID-19 patients admitted during the same months as COVID-19 patients, but these latter estimates were less precise owing to smaller sample size and expected to be more prone to disease misclassification.

### Informing temporal cross-context comparison: characteristics of ICU patients with COVID-19 and non-COVID-19 respiratory conditions, by date of ICU admission

Sociodemographic factors varied more by month of admission among COVID-19 vs. non-COVID-19 patients (**Supp Figure 1**). For example, among COVID-19 patients, mean age was 59.9 years in March 2020 which declined substantially post-Dec 2020 (when UK vaccination programmes began) to 48.9 years in July 2021. Ethnic group proportions also varied more over time among COVID-19 vs. non-COVID-19 patients, with 65.8% of COVID-19 patients of a white ethnic group in March 2020, fluctuating mostly with Asian ethnic group over time, e.g., 38% Asian in May 2021. In contrast, non-COVID-19 patients were more consistently mostly of a white ethnic group over time. Deprivation also varied more over time among COVID-19 patients, with 17% in March 2020 being of the least deprived quintile (10.5% by July 2021; **Supp Figure 2**). The proportion of patients who were of the most deprived quintile increased greatly over time, from 21.8% in March 2020 to a peak of 52% in Aug 2020; a notable increase was seen between Dec 2020 (26.7%; time of vaccine roll-out) and July 2021 (37.2%). Mean BMI varied more over time among COVID-19 patients and showed a modest increase, starting at 29.9 kg/m^2^ in March 2020 and ending at 31.7 kg/m^2^ in July 2021. BMI was relatively stable over time among non-COVID-19 patients. BMI categories also varied more over time among COVID-19 vs. non-COVID-19 patients, e.g., class 3+ obesity was rarest in March 2020 among COVID-19 patients (7.7%), which increased thereafter, ending at 12.8% in July 2021. Comorbidities of COVID-19 patients did not vary more over time than for non-COVID-19 patients, except for prior severe illness. Substantial declines over time were seen in acute severity and number of advanced respiratory support days received among COVID-19 patients, but not among non-COVID-19 patients (**Supp Figure 3-4**). The method of recording BMI varied more over time for COVID-19 patients, with peak winter months showing the highest likelihood of height and weight values being estimated rather than measured (**Supp Figure 4**).

### Informing temporal cross-context comparison: Associations of confounding/selection factors with BMI and mortality among ICU patients with COVID-19 and non-COVID-19 respiratory conditions, by date of ICU admission

Among COVID-19 patients, the associations of confounding/selection factors with BMI often differed by admission date, mostly in terms of magnitude, not direction (**Supp Table 2**). Non-white (vs. white) ethnic groups were associated with lower BMI, deprivation with higher BMI, and most comorbidities with lower BMI except for pre-ICU dependency and comorbid respiratory disease which were mostly associated with higher BMI across dates (e.g., 3.61 kg/m^2^, 95% CI=1.35, 5.87 for comorbid respiratory disease during Feb-April 2021). Among non-COVID-19 patients, the associations of confounding/selection factors with BMI often differed in magnitude across dates, with Asian ethnic group most associated with varying degrees of lower BMI, and with deprivation associated with varying degrees of higher BMI (**Supp Table 3**). Most comorbidities were associated with lower BMI, with the strongest associations being for liver disease (e.g., -3.04 kg/m^2^, 95% CI=-4.64, -1.45 in Aug-Oct 2018).

Among COVID-19 patients, the associations of confounding/selection factors with mortality often differed, again mostly in magnitude, by admission date (**Supp Table 4**). Notably, Black ethnic group was associated with higher mortality only within the first admissions period (Feb-April 2020, HR=1.29, 95% CI=1.15, 1.44); this appeared null during each period thereafter. Most comorbidities were associated with higher mortality across periods, with the largest magnitudes seen with liver disease. Among non-COVID-19 patients, confounding/selection factors were more weakly associated with mortality across periods (**Supp Table 5**). Notably, point estimates indicated that Black ethnic group was associated with lower mortality in each period, with the strongest association in May-July 2018 (HR=0.48, 95% CI=0.26, 0.90). There was weak evidence of association of deprivation with mortality across periods, whilst comorbidities tended to be associated with higher mortality in each period.

### Temporal cross-context comparison: Associations of BMI with mortality among ICU patients with COVID-19 and non-COVID-19 respiratory conditions, by date of ICU admission

Among COVID-19 patients admitted to ICU, higher BMI (per SD) was associated with higher mortality (HR=1.10, 95% CI=1.06, 1.15) during Feb-April 2020 (**Table 3**); this association weakened in May-July 2020 and thereafter with a small positive association during the most populous period of Nov-Jan 2021 (HR=1.04, 95% CI=1.01, 1.07; BMI-date interaction P=0.03). Compared with normal-weight patients, mortality was highest for patients with class 3+ obesity in Feb-April 2020 (HR=1.49, 95% CI=1.27, 1.73), reducing thereafter. A small positive gradient in risk associated with overweight and classes 1 and 2 obesity (vs. normal-weight) was seen only during Feb-April 2020, with HRs for these BMI groups <1 or null across subsequent periods. Being underweight was associated with higher mortality in each period except May-July 2020, but estimates were imprecise given the rarity of underweight COVID-19 patients.

**Table 3.** Temporal cross-context comparison: associations of BMI with 30-day all-cause mortality among ICU patients with COVID-19 and non-COVID-19 respiratory conditions, by admission date

| **HR (95% CI) for mortality, adjusted for sex, age, ethnic group, deprivation, and geographical region** |  |  |  |  |  |  |  |
| --- | --- | --- | --- | --- | --- | --- | --- |
| **<u>COVID-19 patients</u>** |  |  |  |  |  |  |  |
|  | **Feb-April 2020**<br>N=8,054 | **May-July 2020**<br>N=1,470 | **Aug-Oct 2020**<br>N=2,878 | **Nov-Jan 2021**<br>N=15,418 | **Feb-April 2021**<br>N=4,061 | **May-Aug 2021**<br>N=1,929 |  |
| **Adiposity (BMI)** |  |  |  |  |  |  | **P-interaction** |
| *Per SD higher BMI* | *1.10 (1.06, 1.15)* | *0.94 (0.84, 1.05)* | *1.05 (0.97, 1.14)* | *1.04 (1.01, 1.07)* | *1.01 (0.95, 1.07)* | *1.07 (0.96, 1.20)* | *0.03* |
| Underweight | 1.29 (0.82, 2.04) | 0.83 (0.43, 1.59) | 1.02 (0.50, 2.08) | 1.23 (0.87, 1.73) | 1.04 (0.65, 1.68) | 1.92 (1.00, 3.69) |  |
| Normal-weight | 1.00 (reference) | 1.00 (reference) | 1.00 (reference) | 1.00 (reference) | 1.00 (reference) | 1.00 (reference) |  |
| Overweight | 1.07 (0.97, 1.17) | 0.91 (0.72, 1.16) | 1.10 (0.93, 1.30) | 1.00 (0.93, 1.07) | 0.88 (0.75, 1.02) | 0.87 (0.64, 1.20) |  |
| Obesity 1 | 1.03 (0.93, 1.15) | 0.83 (0.63, 1.11) | 0.93 (0.76, 1.13) | 0.97 (0.89, 1.05) | 0.86 (0.73, 1.01) | 0.84 (0.59, 1.18) |  |
| Obesity 2 | 1.13 (0.98, 1.30) | 0.79 (0.55, 1.15) | 1.04 (0.82, 1.32) | 0.95 (0.86, 1.05) | 0.73 (0.60, 0.89) | 1.19 (0.80, 1.75) |  |
| Obesity 3+ | 1.49 (1.27, 1.73) | 0.86 (0.57, 1.30) | 1.15 (0.88, 1.50) | 1.17 (1.05, 1.30) | 1.03 (0.84, 1.27) | 1.17 (0.76, 1.81) |  |
| **<u>Non-COVID-19 patients</u>** |  |  |  |  |  |  |  |
|  | **Feb-April 2018**<br>N=4,587 | **May-July 2018**<br>N=3,266 | **Aug-Oct 2018**<br>N=3,128 | **Nov-Jan 2019**<br>N=5,236 | **Feb-April 2019**<br>N=4,432 | **May-Aug 2019**<br>N=4,230 |  |
| **Adiposity (BMI)** |  |  |  |  |  |  | **P-interaction** |
| *Per SD higher BMI* | *0.81 (0.75, 0.88)* | *0.86 (0.79, 0.94)* | *0.79 (0.72, 0.87)* | *0.94 (0.88, 1.00)* | *0.84 (0.78, 0.91)* | *0.76 (0.71, 0.83)* | *0.001* |
| Underweight | 1.60 (1.26, 2.04) | 1.66 (1.24, 2.23) | 1.45 (1.06, 1.97) | 1.50 (1.18, 1.90) | 1.23 (0.94, 1.62) | 1.72 (1.35, 2.18) |  |
| Normal-weight | 1.00 (reference) | 1.00 (reference) | 1.00 (reference) | 1.00 (reference) | 1.00 (reference) | 1.00 (reference) |  |
| Overweight | 0.82 (0.71, 0.95) | 0.84 (0.71, 1.00) | 0.72 (0.60, 0.86) | 0.96 (0.83, 1.10) | 0.86 (0.74, 1.00) | 0.77 (0.66, 0.90) |  |
| Obesity 1 | 0.76 (0.62, 0.91) | 0.62 (0.48, 0.80) | 0.67 (0.52, 0.85) | 0.85 (0.71, 1.02) | 0.73 (0.59, 0.89) | 0.71 (0.58, 0.88) |  |
| Obesity 2 | 0.60 (0.44, 0.81) | 0.76 (0.54, 1.06) | 0.86 (0.62, 1.20) | 0.85 (0.67, 1.09) | 0.76 (0.57, 1.01) | 0.57 (0.41, 0.78) |  |
| Obesity 3+ | 0.60 (0.43, 0.85) | 1.00 (0.72, 1.39) | 0.59 (0.39, 0.88) | 0.98 (0.75, 1.28) | 0.60 (0.43, 0.84) | 0.64 (0.46, 0.89) |  |

Among non-COVID-19 patients admitted to ICU, higher BMI (per SD) was associated with lower mortality across all admission dates, e.g., HR=0.81 (95% CI=0.75, 0.88) in Feb-April 2018, but with varying strength of association (BMI-date interaction P=0.001). Mortality was highest among underweight patients across all periods, e.g., HR=1.60 (95% CI=1.26, 2.04) during Feb-April 2018. In the same period, mortality was lower with overweight and each obesity group vs. normal-weight, e.g., HR=0.60 (95% CI=0.43, 0.85) for class 3+ obesity. This lower mortality with class 3+ obesity was seen in most periods except for May-July 2018 and Nov-Jan 2019 when mortality did not differ from normal-weight.

### Informing geographical cross-context comparison: characteristics of ICU patients with COVID-19 and non-COVID-19 respiratory conditions, by region of ICU admission

Age and sex appeared similar across regions of admission in both patient groups, whereas proportions of white and Asian ethnic groups varied more across regions among COVID-19 patients (Asian ethnic group being most common in London and least common in Northern Ireland) (**Supp Figure 5-6**). Deprivation varied across regions to a similar degree in patient groups, with northern England showing the highest proportion of most deprived patients (42.7%). Mean/median BMI varied more by region among COVID-19 vs. non-COVID-19 patients, with the lowest BMI in London and the highest BMI in Northern Ireland (**Supp Figure 7**). BMI category proportions also varied more across regions among COVID-19 patients, notably for normal-weight, overweight, and class 3+ obesity (the latter being 7.6% in London and 14.9% in Northern Ireland). Most comorbidities varied similarly across regions among COVID-19 and non-COVID-19 patients, except for greater variability seen for any past severe illness and renal disease among non-COVID-19 patients (**Supp Figure 8**). Notably, the number of advanced respiratory support days varied more across regions among COVID-19 patients, this being lowest in northern England (**Supp Figure 9-10**). Lastly, regional differences in BMI recording practices were similar for COVID-19 and non-COVID-19 patients, with northern England having the most, and Northern Ireland having the least, direct measures of height and weight.

### Informing geographical cross-context comparison: Associations of confounding/selection factors with BMI and mortality among ICU patients with COVID-19 and non-COVID-19 respiratory conditions, by region of ICU admission

Among COVID-19 patients, the existence and magnitude of associations of ethnic and deprivation group with BMI varied by region, e.g., Asian ethnic group was most associated with lower BMI in South England (excluding London) and Northern Ireland (**Supp Table 6**). The magnitude of associations of comorbidities with BMI varied across regions; most of these associations were negative, except for prior dependency which was positively associated with BMI. Among non-COVID-19 patients, there was weaker evidence of associations of confounding/selection factors with BMI varying across regions (**Supp Table 7**).

Among COVID-19 patients, there was weak evidence that the associations of ethnic and deprivation groups with mortality varied by region (**Supp Table 8**). There was stronger evidence that associations of comorbidities with mortality varied by region, mostly in terms of association magnitude. For example, prior severe illness was more strongly associated with higher mortality in Northern Ireland (HR=1.98, 95% CI=1.32, 2.98) than in London (HR=1.18, 95% CI=1.06, 1.31). Among non-COVID-19 patients, there was generally weak evidence of association of sociodemographic factors with mortality across regions, except for Black ethnic group which was often associated with lower mortality, e.g., HR=0.78 (95% CI=0.61, 1.01) in London (**Supp Table 9**). There was stronger evidence that associations of comorbidities with mortality varied across regions, most notably for end-stage renal disease which was associated with lower mortality in South England (HR=0.33, 95% CI=0.15, 0.74) but higher mortality in Northern Ireland (HR=6.49, 95% CI=2.56, 16.48).

### Geographical cross-context comparison: Associations of BMI with mortality among ICU patients with COVID-19 and non-COVID-19 respiratory conditions, by region of ICU admission

Among COVID-19 patients admitted to ICU, the association of BMI (per SD higher) with higher mortality did not vary across regions (BMI-region interaction P=0.75; **Table 4**). Compared with normal-weight, mortality was most consistently elevated with class 3+ obesity across regions, with the highest risk in North England (HR=1.26; 95% CI=1.11, 1.44) and London (HR=1.23; 95% CI=1.04, 1.46). Neither overweight nor class 1 or 2 obesity were clearly associated with mortality across regions, with the mortality HR for class 2 obesity imprecisely raised in Northern Ireland, at 1.41 (95% CI=0.85, 2.32).

**Table 4.**
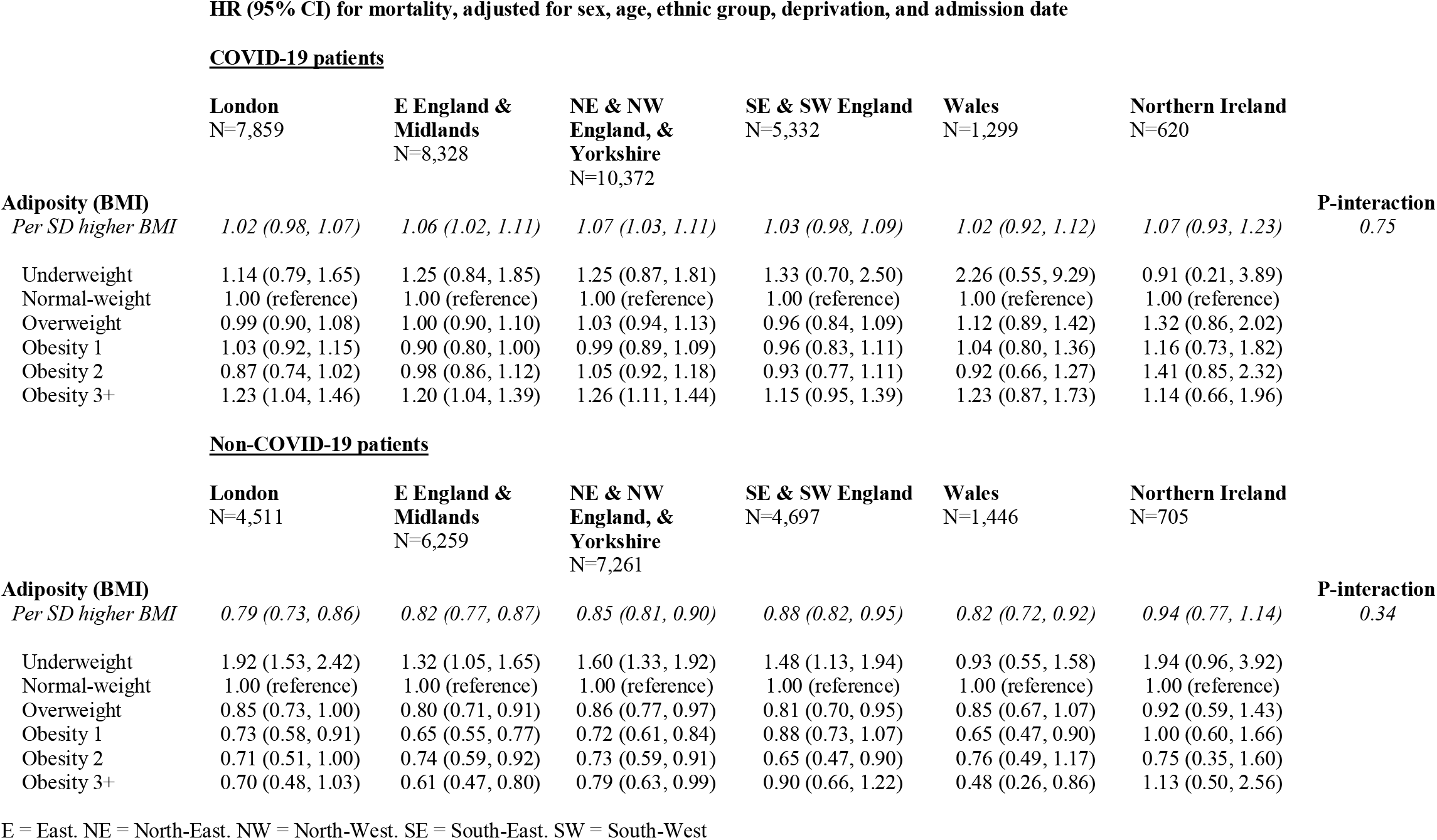
Geographical cross-context comparison: associations of BMI with 30-day all-cause mortality among ICU patients with COVID-19 (5 Feb 2020 to 1 Aug 2021) and non-COVID-19 respiratory conditions (1 Feb 2018 to 31 Aug 2019), by geographical region

| **HR (95% CI) for mortality, adjusted for sex, age, ethnic group, deprivation, and admission date** |  |  |  |  |  |  |  |
| --- | --- | --- | --- | --- | --- | --- | --- |
| **<u>COVID-19 patients</u>** |  |  |  |  |  |  |  |
|  | **London**<br>N=7,859 | **E England & Midlands**<br>N=8,328 | **NE & NW England, & Yorkshire**<br>N=10,372 | **SE & SW England**<br>N=5,332 | **Wales**<br>N=1,299 | **Northern Ireland**<br>N=620 |  |
| **Adiposity (BMI)** |  |  |  |  |  |  | **P-interaction** |
| *Per SD higher BMI* | *1.02 (0.98, 1.07)* | *1.06 (1.02, 1.11)* | *1.07 (1.03, 1.11)* | *1.03 (0.98, 1.09)* | *1.02 (0.92, 1.12)* | *1.07 (0.93, 1.23)* | *0.75* |
| Underweight | 1.14 (0.79, 1.65) | 1.25 (0.84, 1.85) | 1.25 (0.87, 1.81) | 1.33 (0.70, 2.50) | 2.26 (0.55, 9.29) | 0.91 (0.21, 3.89) |  |
| Normal-weight | 1.00 (reference) | 1.00 (reference) | 1.00 (reference) | 1.00 (reference) | 1.00 (reference) | 1.00 (reference) |  |
| Overweight | 0.99 (0.90, 1.08) | 1.00 (0.90, 1.10) | 1.03 (0.94, 1.13) | 0.96 (0.84, 1.09) | 1.12 (0.89, 1.42) | 1.32 (0.86, 2.02) |  |
| Obesity 1 | 1.03 (0.92, 1.15) | 0.90 (0.80, 1.00) | 0.99 (0.89, 1.09) | 0.96 (0.83, 1.11) | 1.04 (0.80, 1.36) | 1.16 (0.73, 1.82) |  |
| Obesity 2 | 0.87 (0.74, 1.02) | 0.98 (0.86, 1.12) | 1.05 (0.92, 1.18) | 0.93 (0.77, 1.11) | 0.92 (0.66, 1.27) | 1.41 (0.85, 2.32) |  |
| Obesity 3+ | 1.23 (1.04, 1.46) | 1.20 (1.04, 1.39) | 1.26 (1.11, 1.44) | 1.15 (0.95, 1.39) | 1.23 (0.87, 1.73) | 1.14 (0.66, 1.96) |  |
| **<u>Non-COVID-19 patients</u>** |  |  |  |  |  |  |  |
|  | **London**<br>N=4,511 | **E England & Midlands**<br>N=6,259 | **NE & NW England, & Yorkshire**<br>N=7,261 | **SE & SW England**<br>N=4,697 | **Wales**<br>N=1,446 | **Northern Ireland**<br>N=705 |  |
| **Adiposity (BMI)** |  |  |  |  |  |  | **P-interaction** |
| *Per SD higher BMI* | *0.79 (0.73, 0.86)* | *0.82 (0.77, 0.87)* | *0.85 (0.81, 0.90)* | *0.88 (0.82, 0.95)* | *0.82 (0.72, 0.92)* | *0.94 (0.77, 1.14)* | *0.34* |
| Underweight | 1.92 (1.53, 2.42) | 1.32 (1.05, 1.65) | 1.60 (1.33, 1.92) | 1.48 (1.13, 1.94) | 0.93 (0.55, 1.58) | 1.94 (0.96, 3.92) |  |
| Normal-weight | 1.00 (reference) | 1.00 (reference) | 1.00 (reference) | 1.00 (reference) | 1.00 (reference) | 1.00 (reference) |  |
| Overweight | 0.85 (0.73, 1.00) | 0.80 (0.71, 0.91) | 0.86 (0.77, 0.97) | 0.81 (0.70, 0.95) | 0.85 (0.67, 1.07) | 0.92 (0.59, 1.43) |  |
| Obesity 1 | 0.73 (0.58, 0.91) | 0.65 (0.55, 0.77) | 0.72 (0.61, 0.84) | 0.88 (0.73, 1.07) | 0.65 (0.47, 0.90) | 1.00 (0.60, 1.66) |  |
| Obesity 2 | 0.71 (0.51, 1.00) | 0.74 (0.59, 0.92) | 0.73 (0.59, 0.91) | 0.65 (0.47, 0.90) | 0.76 (0.49, 1.17) | 0.75 (0.35, 1.60) |  |
| Obesity 3+ | 0.70 (0.48, 1.03) | 0.61 (0.47, 0.80) | 0.79 (0.63, 0.99) | 0.90 (0.66, 1.22) | 0.48 (0.26, 0.86) | 1.13 (0.50, 2.56) |  |

E = East. NE = North-East. NW = North-West. SE = South-East. SW = South-West

Among non-COVID-19 patients admitted to ICU, the association of BMI (per SD higher) with lower mortality did not vary across regions, e.g., HR=0.79 (95% CI=0.73, 0.86) in London (BMI-region interaction P=0.34; **Table 4**). Compared with being normal-weight, only being underweight was associated with higher mortality across regions, which differed in association magnitude (e.g., HR=1.92; 95% CI=1.53, 2.42 in London and HR=1.32; 95% CI=1.05, 1.65 in East England and Midlands). Across most regions, mortality was lower for each BMI group above normal-weight, with class 3+ obesity often carrying the lowest risk, e.g. at HR=0.61 (95% CI=0.47, 0.80) in East England and Midlands and HR=0.48 (95% CI=0.26, 0.86) in Wales.

## Discussion

We aimed in this study to assess the causality of adiposity for mortality among patients severely ill with COVID-19 and non-COVID-19 respiratory conditions using a cross-context comparison approach with nationally representative ICU data in the UK. Consistent adiposity-mortality associations despite varying confounding/selection would increase confidence in causality. Our results suggest that higher adiposity, primarily extreme obesity, is associated with higher mortality among patients admitted to ICU with COVID-19, but lower mortality among patients admitted with non-COVID-19 respiratory conditions. These associations appear vulnerable to confounding/selection bias in both patient groups, questioning the existence or stability of causal effects. Among COVID-19 patients, unfavourable obesity-mortality associations differ substantially by ICU admission date. Among non-COVID-19 respiratory patients, favourable obesity-mortality associations may reflect comorbidity-induced weight loss.

The two contexts examined here were date and geographical region of ICU admission. These were chosen because the prevalence/level of confounding and selection factors relevant to adiposity-mortality effects differ by time and geography among ICU patients with COVID-19 (20), and thus the effects of those confounders/selectors on adiposity/mortality might also differ amongst them, although this had not been previously examined. Such context-varying bias is expected to be most influential for COVID-19 given rapid changes in its clinical/public management, whereas context-stable bias is expected to be most influential for non-COVID-19 respiratory conditions which was assessed here by comparing characteristics between patient groups. Our results suggest that the associations of confounding/selection factors with BMI and mortality varied mostly by date (not region) of ICU admission, and particularly for COVID-19. For example, among COVID-19 patients, Black ethnic group was associated with different degrees of lower BMI across admission dates, and with higher mortality only during Feb-April 2020. This diminishing association with mortality over time has also been seen in the UK general population outside of ICU settings (24). Deprivation showed little variation in its association with BMI or mortality by date, but variation was greater for several comorbidity indicators including severe liver disease. Notably, higher BMI was not consistently associated with higher mortality across admission dates among COVID-19 patients, with ∼50% higher mortality seen for extreme obesity only within the earliest period of Feb-April 2020. More consistent (negative) obesity-mortality associations were seen among non-COVID-19 patients. This consistency among non-COVID-19 patients does not necessarily support causality in that group, however, as this could still reflect stable forms of selection bias such as reverse causation. This was supported by comparisons of patient group characteristics which indicated over-admissions to ICU of leaner patients with more comorbidities who experience higher mortality, or comorbidity-induced weight loss/cachexia.

Adiposity-mortality associations may differ over time because of bias or because of genuine changes in causality, e.g., due to changes in viral variants, vaccines, and therapeutics. COVID-19 vaccines were introduced in the UK in Dec 2020 and uptake was earliest among the oldest and most clinically vulnerable (including both extreme obesity and underweight) – the same populations who present most often to ICU. Vaccines substantially reduce mortality from SARS-CoV-2 infection and COVID-19 (25), and along with improved therapeutics likely explain overall declines in mortality, acute severity, and mean age of patients admitted to ICU with COVID-19 post-2020. Notably, however, our results suggest that adiposity-mortality associations among COVID-19 patients started diminishing in May-July 2020, before vaccines were introduced in Dec 2020. This suggests that vaccines/therapeutics are not the sole reason for variation over time in adiposity-mortality associations among COVID-19 patients, and that confounding/selection bias which also varied over time played a role. For example, collider (selection) bias (17, 26) could have manifested in COVID-19 patients being admitted to ICU at higher BMIs yet with fewer comorbidities over the course of the pandemic, following early evidence on obesity-mortality (5, 15). Our descriptions of patient characteristics over time do suggest increased mean/median BMI (notably overweight/mild obesity) of patients admitted to ICU with COVID-19 from July 2020, while the prevalence of past severe illness declined/fluctuated over the same period and was lowest while mean BMI was highest in Feb-April 2020. These negative BMI-comorbidity associations among COVID-19 patients selected into ICU could have biased obesity-mortality associations in Feb-April 2020 when obesity appears to raise mortality, and/or in later time periods where obesity appears not to raise mortality. Diminishing obesity-mortality associations over time could also be explained by biologically different immune responses to SARS-CoV-2 for repeated vs. immunologically naïve infections if later admissions included more patients with repeat infections (27-29).

Our results and the results of earlier descriptive reports by ICNARC (20) indicate that obesity, including extreme obesity, is more common within COVID-19 than non-COVID-19 respiratory conditions. It is well known that severe COVID-19 occurs more frequently in patients with more comorbidities in the general population (30), but our results suggest that, within ICU, patients with COVID-19 tend to have fewer comorbidities than patients with non-COVID-19 respiratory conditions. This is despite obesity being more common among COVID-19 patients. This same pattern was also seen in the Netherlands, based on one study using ICU data which compared the adiposity and comorbidity profile of ∼2,600 ICU patients with COVID-19 vs. ∼2,900 with non-COVID-19 viral pneumonia (31). That Dutch study reported the same contrasting pattern of BMI-mortality associations between patient groups: positive among COVID-19 patients and negative among non-COVID-19 respiratory patients. This obesity paradox in ICU is striking and may help reveal the potential for patient selection and reverse causation to bias BMI-mortality associations within severe respiratory disease more broadly – with COVID-19 vs. non-COVID-19 offering another type of cross-context comparison. Pre-existing disease may reduce BMI and raise mortality, and this may explain long-standing observations of higher mortality with underweight and lower mortality with obesity (vs. normal-weight) among respiratory disease patients (10, 11). Given the relatively healthy comorbidity profile of COVID-19 patients, associations between BMI and mortality amongst them may be less subject to reverse causation, and thus inform on the likely causality of BMI for mortality among both COVID-19 and non-COVID-19 patients (if these conditions are clinically similar). This is important given the need for appropriate clinical messaging around obesity during potential dual burdens of COVID-19 and influenza in future. Our results suggest unfavourable obesity-mortality associations among COVID-19 patients (who have fewer comorbidities), in contrast to favourable obesity-mortality associations among non-COVID-19 patients (who have more comorbidities). If comorbidity-induced weight loss is expected to bias associations then these results suggest that obesity may not be protective in either group and that weight loss/maintenance advice applies to both groups.

Over a dozen previous studies examined associations of BMI with mortality within severe COVID-19; most were small scale (N<500), with larger studies suggesting that excess mortality is driven by extreme obesity, where this was examined (32-37). Given the global spread of COVID-19, the totality of studies examining adiposity-mortality associations naturally provides a comparison of these associations across temporal and geographical contexts, but the study designs and methods used differ in many other respects and no previous study directly compared dates or regions using the same analytical strategy. One UK study examined how associations between ethnicity among mortality among COVID-19 patients changed over calendar time, but did not examine BMI (24). Importantly, none of these past studies directly compared the associations of confounding/selection factors with adiposity and mortality across contexts to appraise their impact. Previous studies also tended to statistically adjust for comorbidities in main effect models, which may be an overadjustment and could induce collider bias given that comorbidities can result from adiposity. With cross-context comparisons, however, it is difficult to identify specific confounding or selection factors which underpin any differences in exposure-outcome associations; multiple factors are likely influential, many of which are likely unmeasured and are only proxied by factors which are stratified on. Interpreting results necessarily relies on critical judgement and assessing causality is inherently qualitative.

### Limitations

This study is observational and associations are subject to confounding, selection bias, reverse causation, and measurement error. Cross-context comparisons are intended to interrogate the extent and impact of such biases on exposure-outcome associations but offer incomplete and qualitative assessments. Measurement error may be problematic for adiposity given that this was measured indirectly using BMI which correlates less well with more objective measures of fat mass in severely ill vs. young healthy populations, although the correlation between BMI and abdominal fat area is ∼0.7 among severely ill adults (38, 39). Data are also collected within ‘real world’ ICU settings and are often recorded less accurately than in research-grade clinics. BMI measures here were a mixture of directly measured and estimated values upon ICU admission, and these proportions varied more over time for COVID-19 vs. non-COVID-19 patients (reflecting changing workloads/resources during virus waves). The extent of estimated values was similar between patient groups, however, and these different methods of BMI recording have previously shown consistent associations with ICU mortality (22).

Our study is limited to data which are routinely collected in ICU settings nationally, and thus data on other adiposity measures such as waist circumference and lifestyle factors such as smoking, diet, and physical activity were not available. Smoking history data would be particularly useful for assessing confounding, e.g., where excess mortality with underweight in non-COVID-19 may have been partly confounded by effects of smoking on weight loss and mortality. Comorbid disease indicators were also limited to very severe forms of disease and exclude less severe diseases which are still relevant to mortality, such as type 2 diabetes and other cardiovascular diseases. One Dutch ICU study did, however, record diabetes history in patients with COVID-19 and non-COVID-19 pneumonia and found no difference between groups (∼20% in each) (31).

Lastly, the ICU data used here were representative of adult populations in England, Wales, and Northern Ireland, but likely excluded individuals who were most extremely clinically vulnerable as they would have been shielding during peak stages of the pandemic and thus presenting less than usual to ICU, whereas equivalently extremely vulnerable non-COVID-19 respiratory patients would have presented more readily. Hospital practices were also atypical during COVID-19 surges and many patients severely ill with COVID-19 were likely managed outside of ICU on regular wards due to limited capacity. Such practices would have likely varied more by time than by geography given that national clinical guidance and protocols were rapidly shared across regions of the UK via the NHS during the pandemic.

### Conclusions

Our results based on a cross-context comparison approach with nationally representative ICU data in the UK suggest that higher adiposity, primarily extreme obesity, is associated with higher mortality among patients admitted to ICU with COVID-19, but lower mortality among patients admitted with non-COVID-19 respiratory conditions. These associations appear vulnerable to confounding/selection bias in both patient groups, questioning the existence or stability of causal effects. Among COVID-19 patients, unfavourable obesity-mortality associations differ substantially by admission date. Among non-COVID-19 respiratory patients, favourable obesity-mortality associations may reflect comorbidity-induced weight loss.

## Data Availability

Individual-level data are available via application to ICNARC (managed access).

## Acknowledgements

We thank and respect all those working in critical care units across England, Wales, and Northern Ireland and contributing to the care of patients and, particularly, those responsible for submitting data rapidly and regularly during the COVID-19 epidemic. Additional Intensive Care National Audit & Research Centre Coronavirus Disease 2019 Team Members: Yemi Banjo, Kasia Borowczak, Tom Cousins, Peter Cummins, Keji Dalemo, Robert Darnell, Hanna Demissie, Laura Drikite, Andrew Fleming, Ditte Frederiksen, Sarah Furnell, Abdo Hussein, Abby Koelewyn, Tim Matthews, Izabella Orzechowska, Sam Peters, Alvin Richards-Belle, Tyrone Samuels, and Michelle Saull.

## Notes

**Funding:** JAB, DC, and AH are supported by the Elizabeth Blackwell Institute for Health Research and the Development and Alumni Relations Office, University of Bristol. JAB, DC, AH, KT, and GDS work in a unit funded by the UK MRC (MC_UU_00011/1; MC_UU_00011/3) and the University of Bristol. This publication is the work of the authors and JAB is the guarantor for its contents. The funders had no role in study design, data collection and analysis, decision to publish, or preparation of the manuscript.

**Conflicts of interest:** None to declare.

### Competing Interest Statement

The authors have declared no competing interest.

### Funding Statement

JAB, DC, and AH are supported by the Elizabeth Blackwell Institute for Health Research and the Development and Alumni Relations Office, University of Bristol. JAB, DC, AH, KT, and GDS work in a unit funded by the UK MRC (MC_UU_00011/1; MC_UU_00011/3) and the University of Bristol. This publication is the work of the authors and JAB is the guarantor for its contents. The funders had no role in study design, data collection and analysis, decision to publish, or preparation of the manuscript.

### Author Declarations

Approval for the collection and use of patient-identifiable data without consent in the Case Mix Programme was obtained from the Confidentiality Advisory Group of the Health Research Authority under Section 251 of the NHS Act 2006 (approval number PIAG2-10[f]/2005). All data were pseudonymised (patient identifiers removed) prior to extraction for this research.

