## Supplementary Figures & Tables for "Adiposity and Mortality among Patients Severely Ill with COVID-19 and non-COVID-19 Respiratory Conditions: A Cross-Context Comparison Study in the UK"

**Supplementary Figure 1** Age, sex, and ethnic group profiles of ICU patients with COVID-19 (1 March 2020 to 31 July 2021) and non-COVID-19 respiratory conditions (1 Feb 2018 to 31 Aug 2019), by admission date

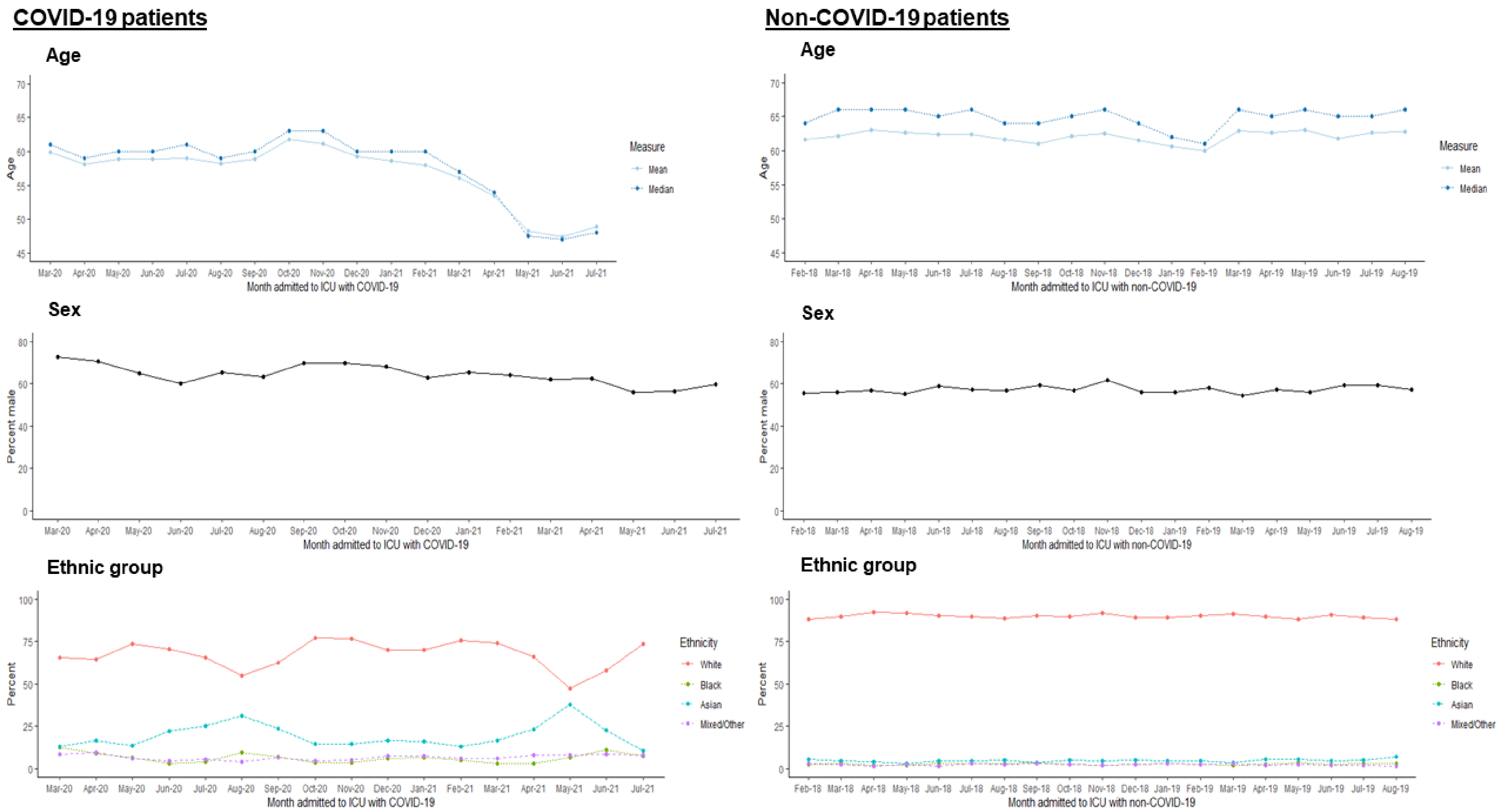


**Note:** Feb 2020 and Aug 2021 are excluded from COVID-19 profiles due to low case counts (N=5 and N=3 respectively)

**Supplementary Figure 2** Deprivation and adiposity profiles of ICU patients with COVID-19 (1 March 2020 to 31 July 2021) and non-COVID-19 respiratory conditions (1 Feb 2018 to 31 Aug 2019), by admission date

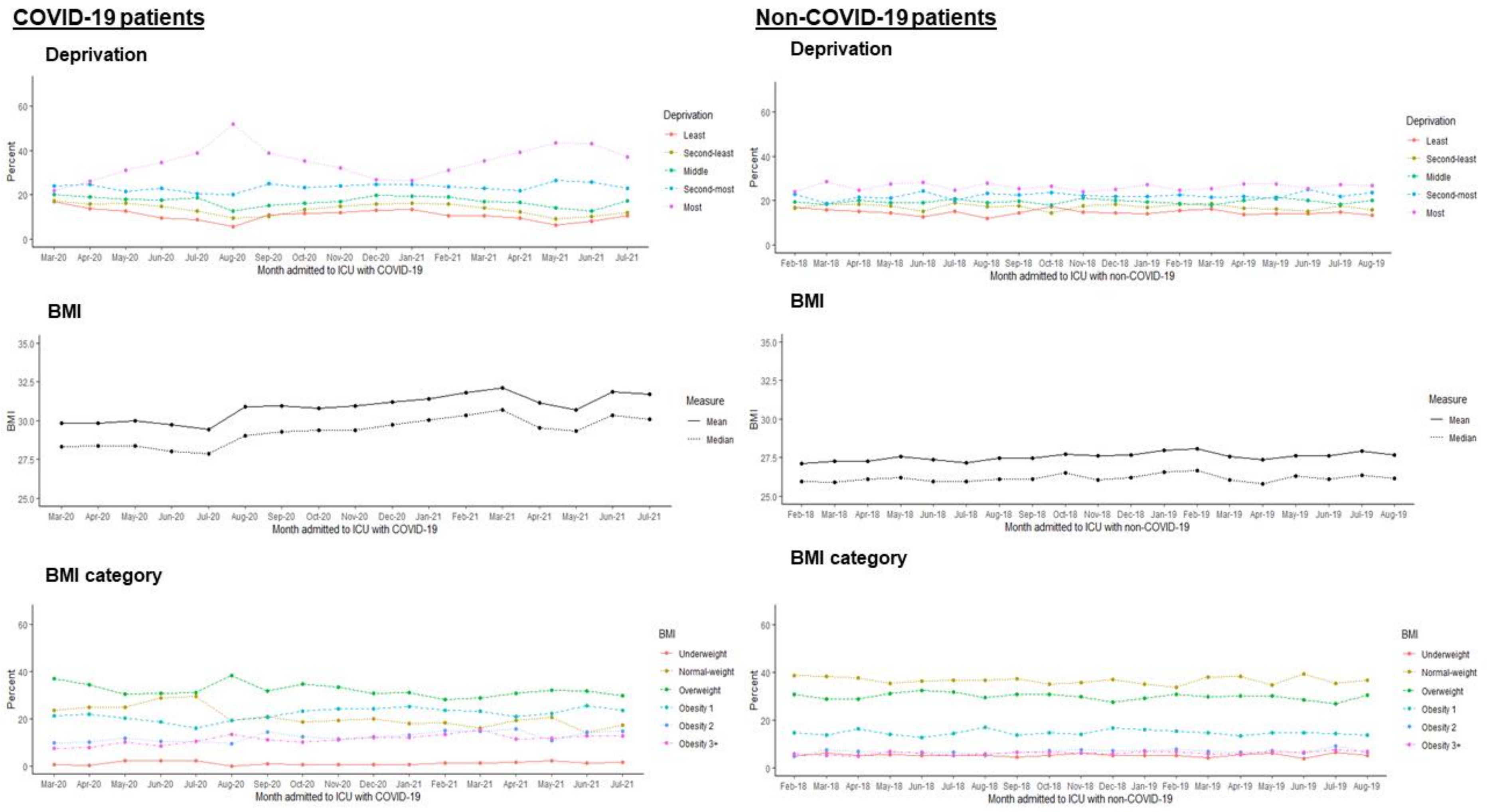


**Note:** Feb 2020 and Aug 2021 are excluded from COVID-19 profiles due to low case counts (N=5 and N=3 respectively)

**Supplementary Figure 3** Dependency, comorbidity, and acute severity profiles of ICU patients with COVID-19 (1 March 2020 to 31 July 2021) and non-COVID-19 respiratory conditions (1 Feb 2018 to 31 Aug 2019), by admission date

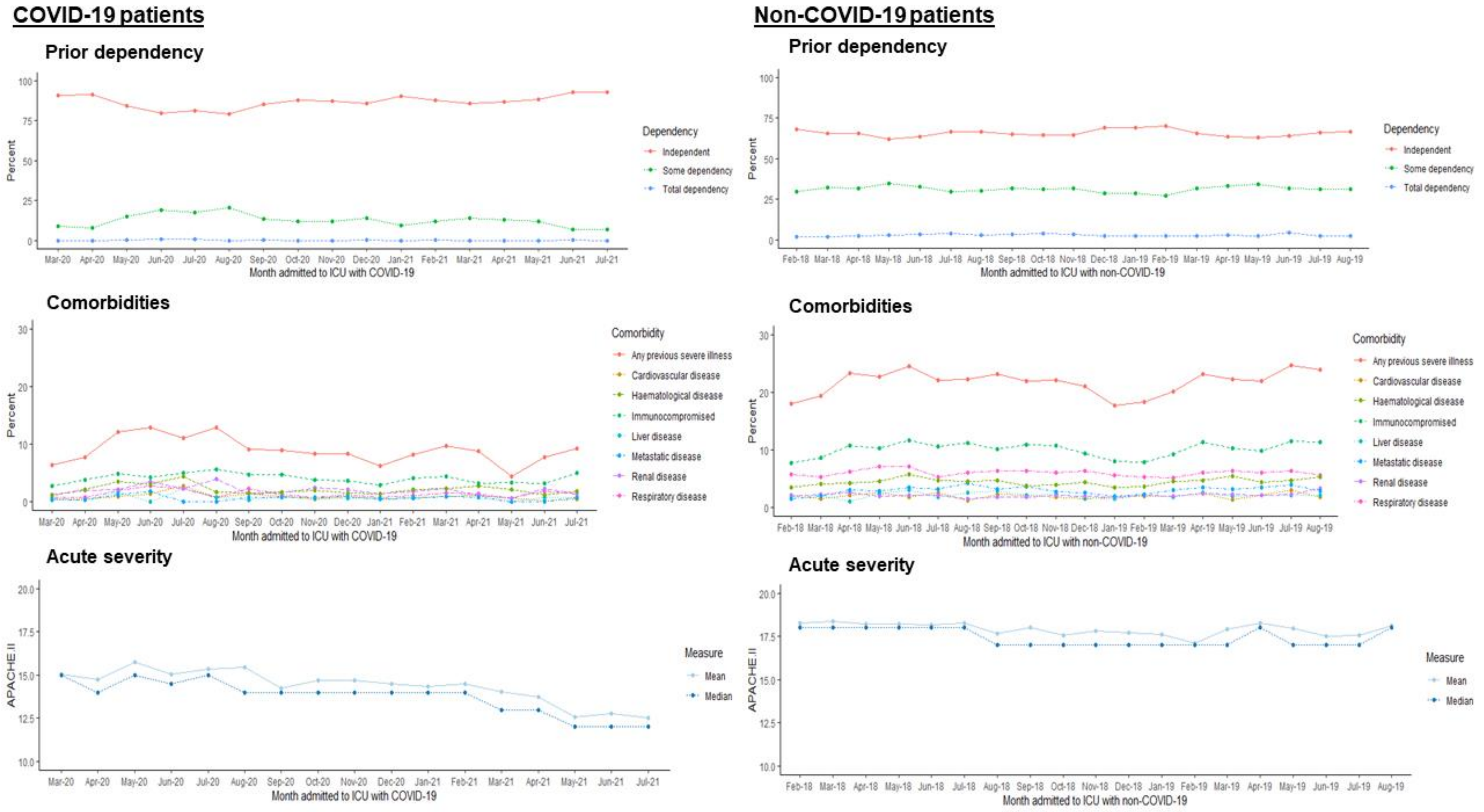


**Note:** Feb 2020 and Aug 2021 are excluded from COVID-19 profiles due to low case counts (N=5 and N=3 respectively)

**Supplementary Figure 4** Respiratory severity and support profiles of ICU patients with COVID-19 (1 March 2020 to 31 July 2021) and non-COVID-19 respiratory conditions (1 Feb 2018 to 31 Aug 2019), by admission date

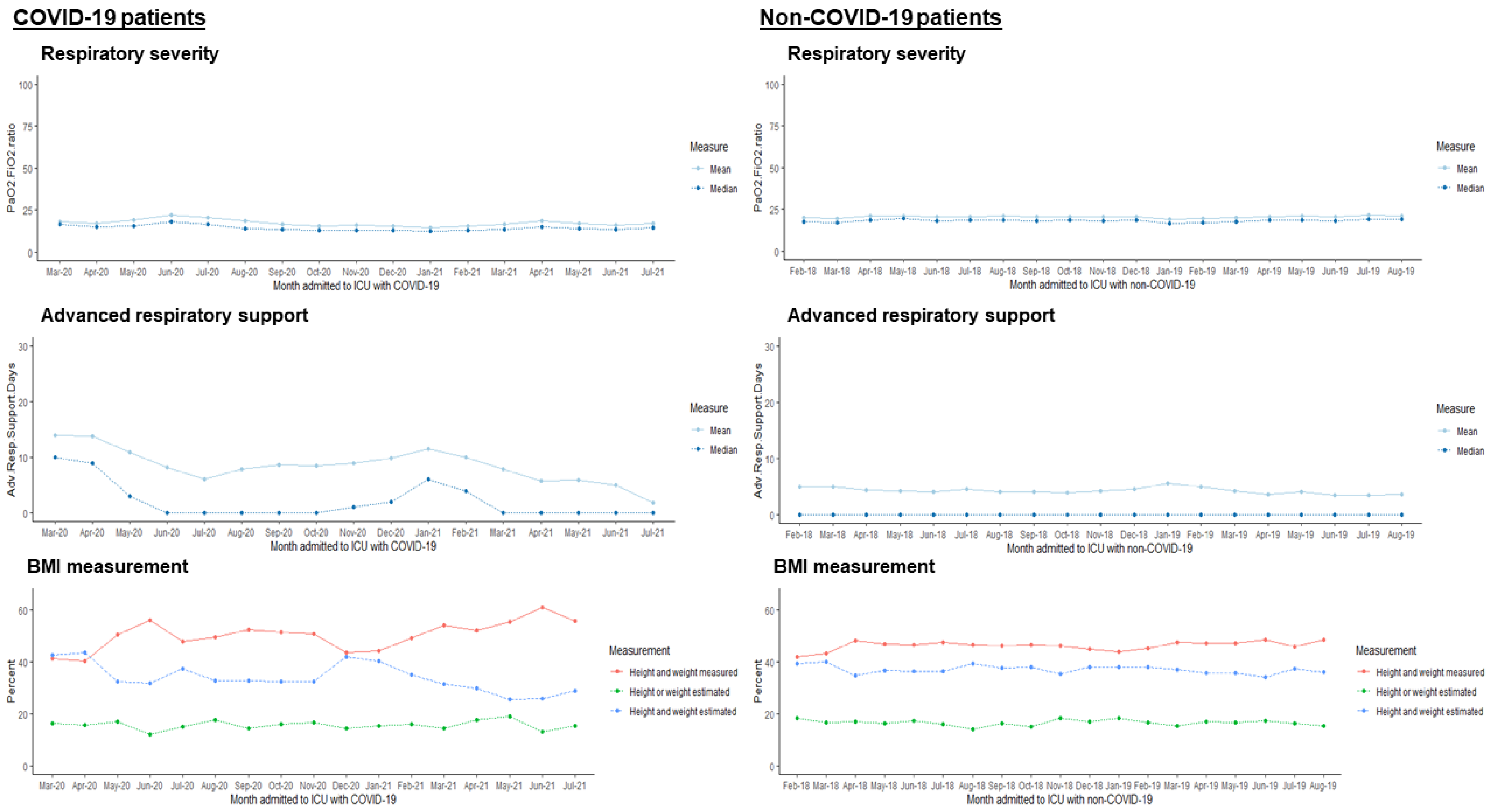


**Note:** Feb 2020 and Aug 2021 are excluded from COVID-19 profiles due to low case counts (N=5 and N=3 respectively)

**Supplementary Figure 5** Age and sex profiles of ICU patients with COVID-19 (5 Feb 2020 to 1 Aug 2021) and non-COVID-19 respiratory conditions (1 Feb 2018 to 31 Aug 2019), by geographical region

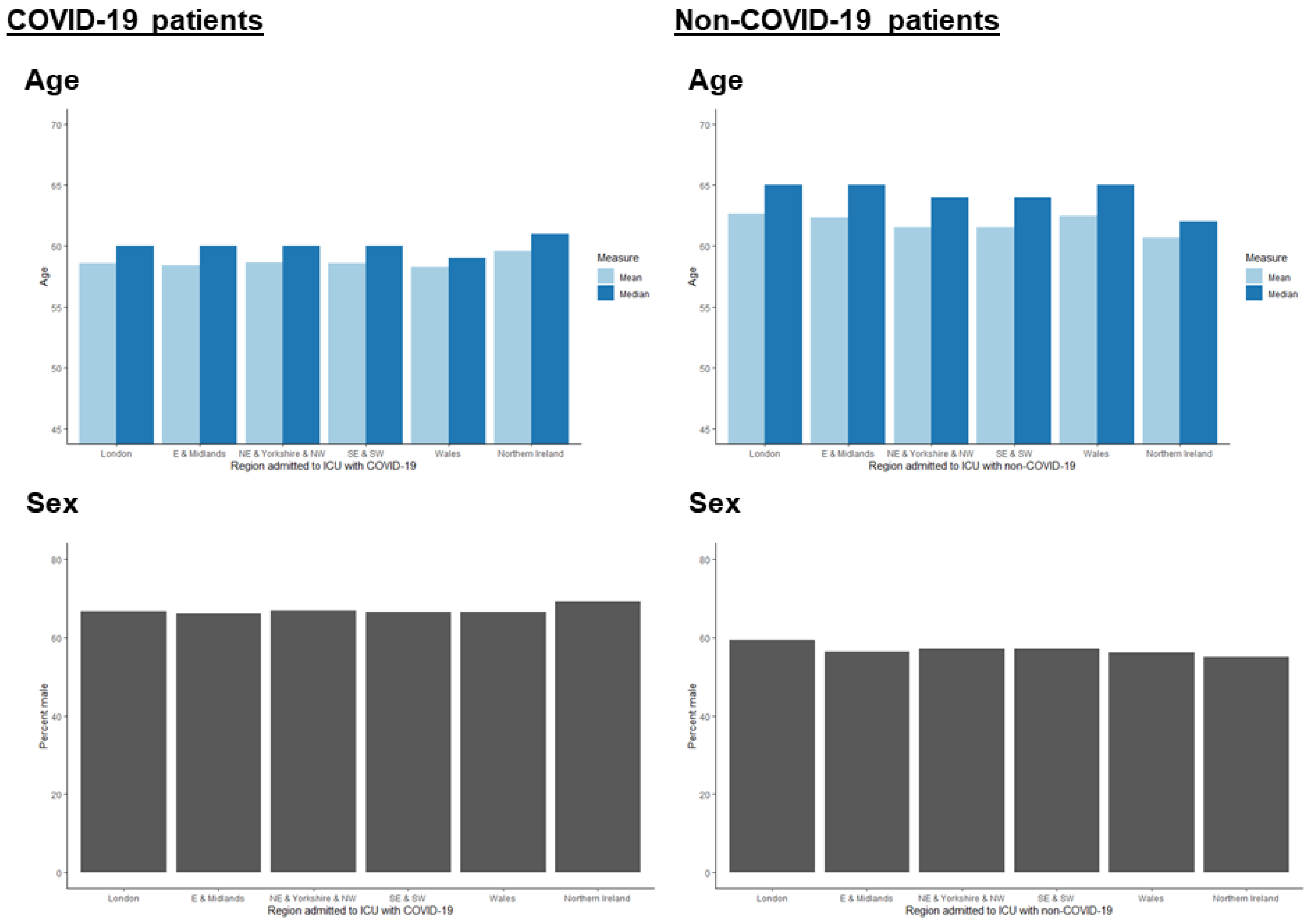


E = East. NE = North-East. NW = North-West. SE = South-East. SW = South-West

**Supplementary Figure 6** Ethnic group and deprivation profiles of ICU patients with COVID-19 (5 Feb 2020 to 1 Aug 2021) and non-COVID-19 respiratory conditions (1 Feb 2018 to 31 Aug 2019), by geographical region

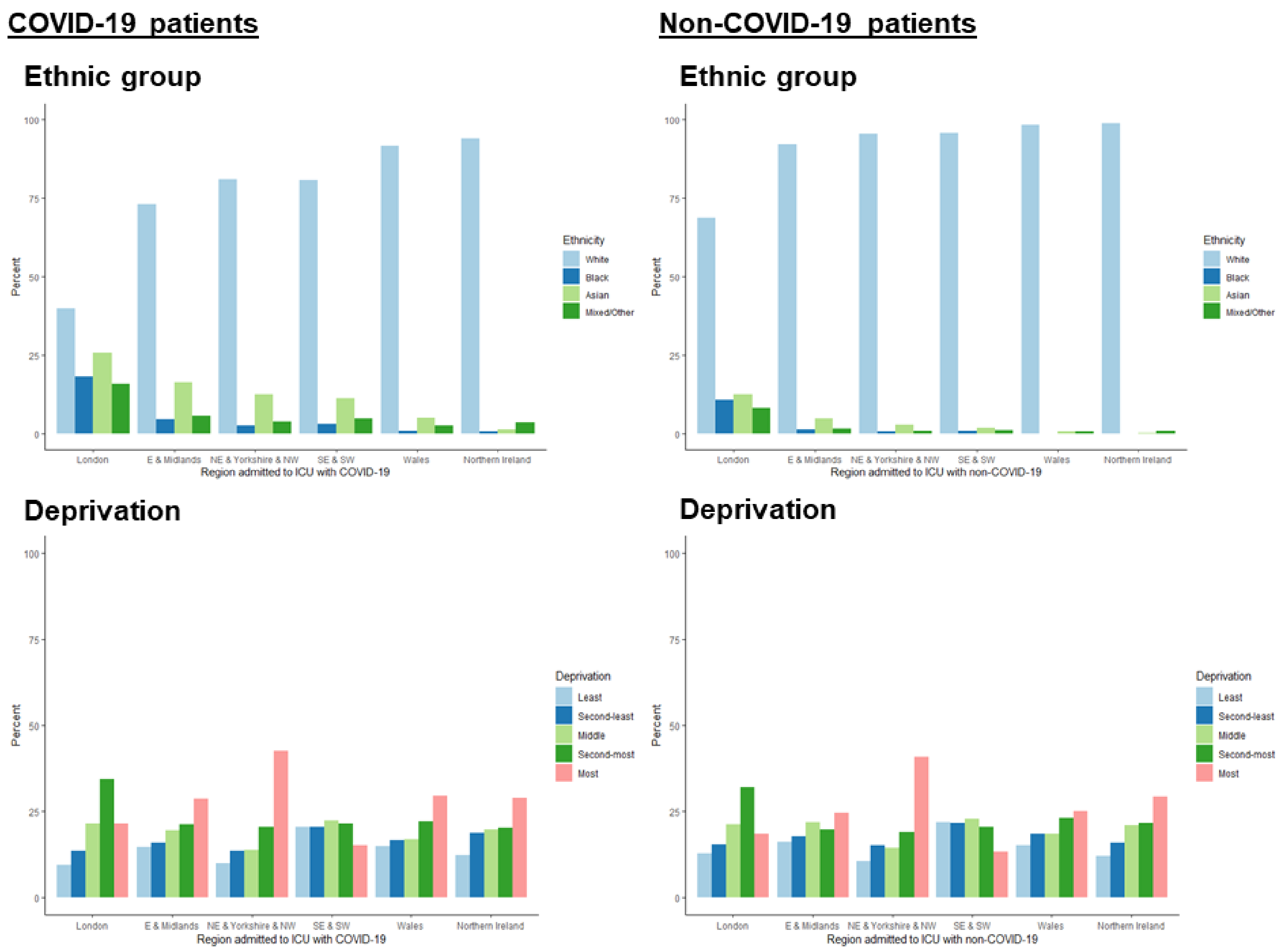


E = East. NE = North-East. NW = North-West. SE = South-East. SW = South-West

**Supplementary Figure 7** Adiposity profiles of ICU patients with COVID-19 (5 Feb 2020 to 1 Aug 2021) and non-COVID-19 respiratory conditions (1 Feb 2018 to 31 Aug 2019), by geographical region

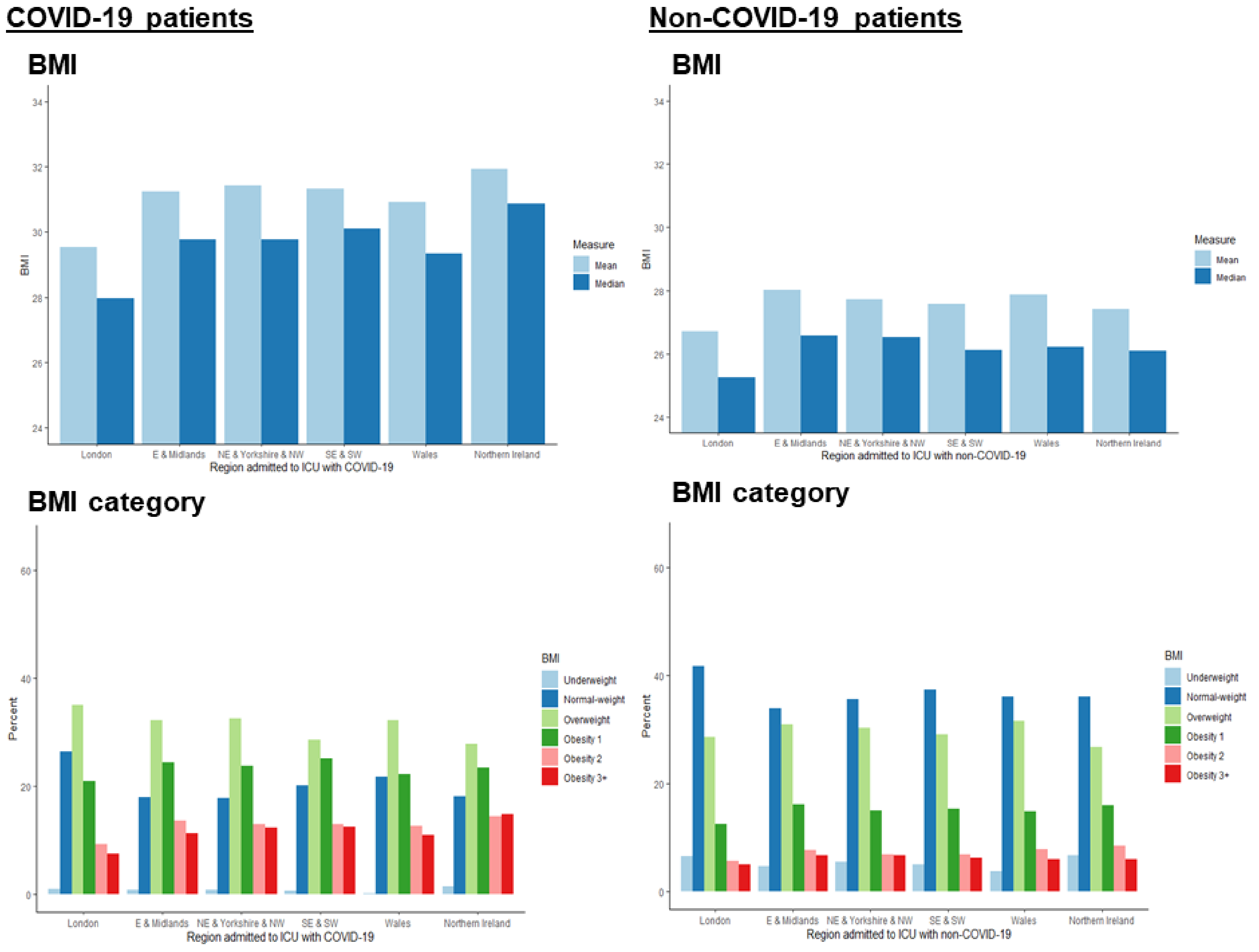


E = East. NE = North-East. NW = North-West. SE = South-East. SW = South-West

**Supplementary Figure 8** Prior dependency and comorbidity profiles of ICU patients with COVID-19 (5 Feb 2020 to 1 Aug 2021) and non-COVID-19 respiratory conditions (1 Feb 2018 to 31 Aug 2019), by geographical region

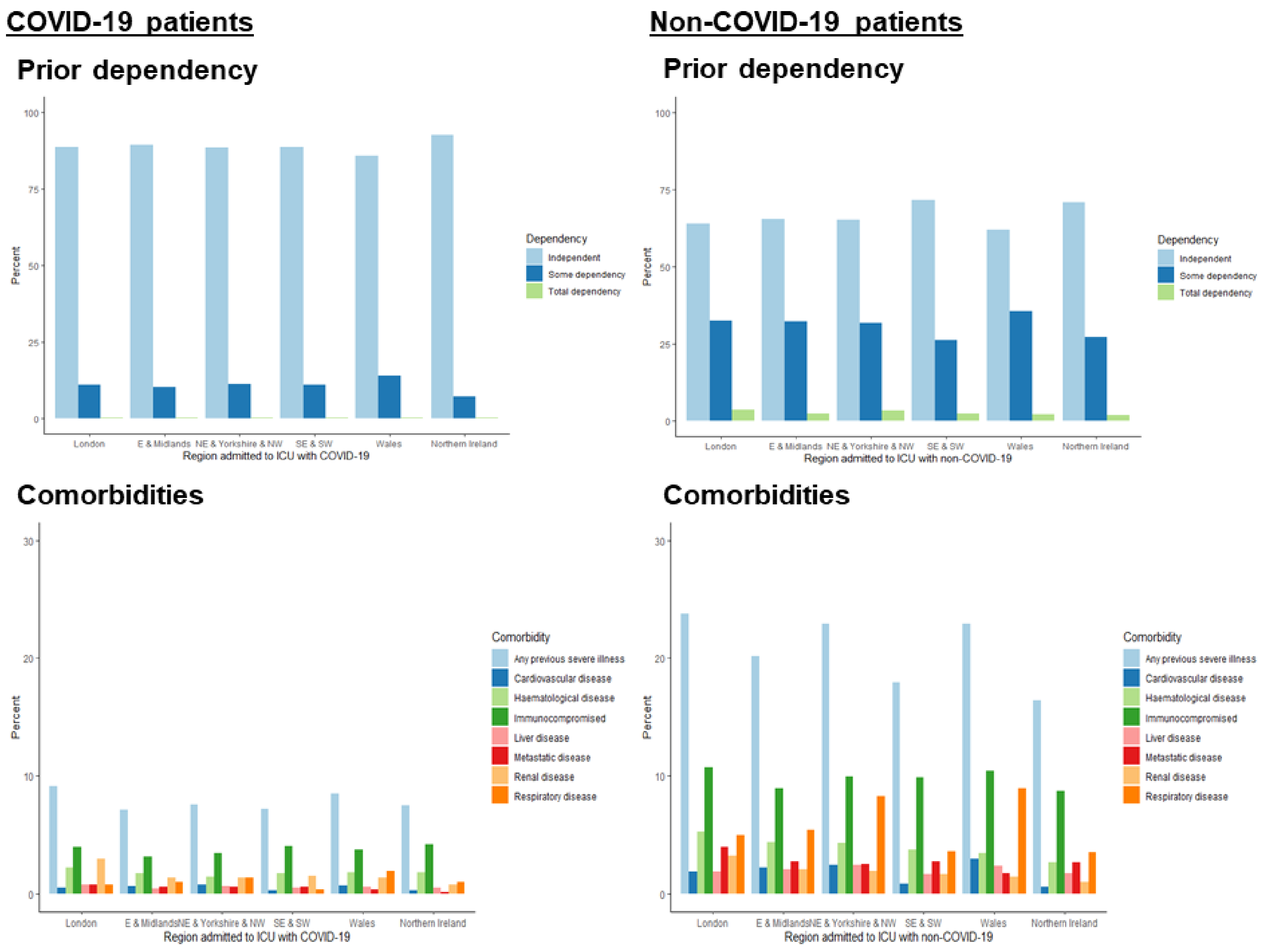


E = East. NE = North-East. NW = North-West. SE = South-East. SW = South-West

**Supplementary Figure 9** Acute and respiratory severity profiles of ICU patients with COVID-19 (5 Feb 2020 to 1 Aug 2021) or non-COVID-19 respiratory conditions (1 Feb 2018 to 31 Aug 2019), by geographical region

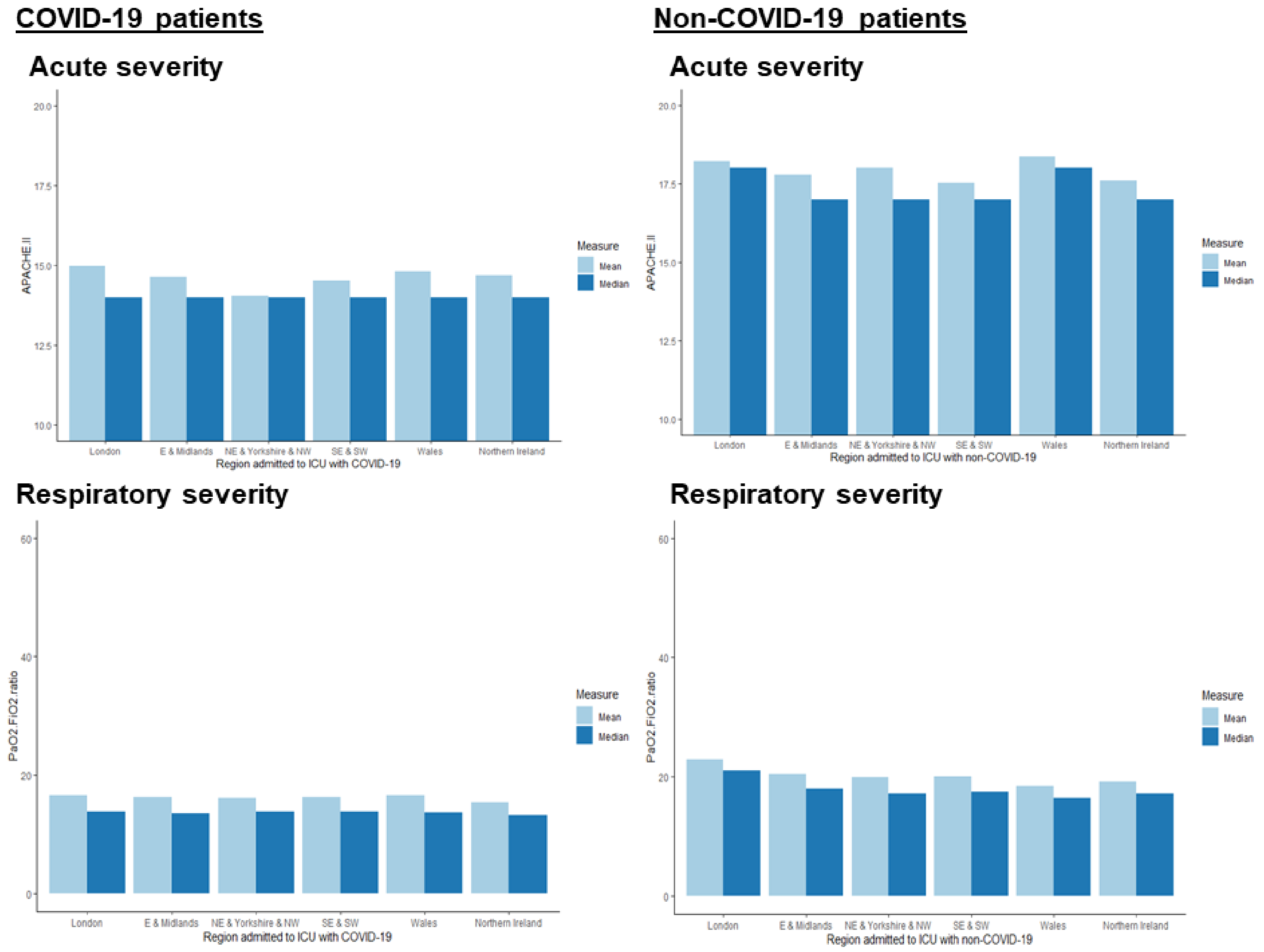


E = East. NE = North-East. NW = North-West. SE = South-East. SW = South-West

**Supplementary Figure 10** Advanced respiratory support and adiposity measurement profiles of ICU patients with COVID-19 (5 Feb 2020 to 1 Aug 2021) or non-COVID-19 respiratory conditions (1 Feb 2018 to 31 Aug 2019), by geographical region.

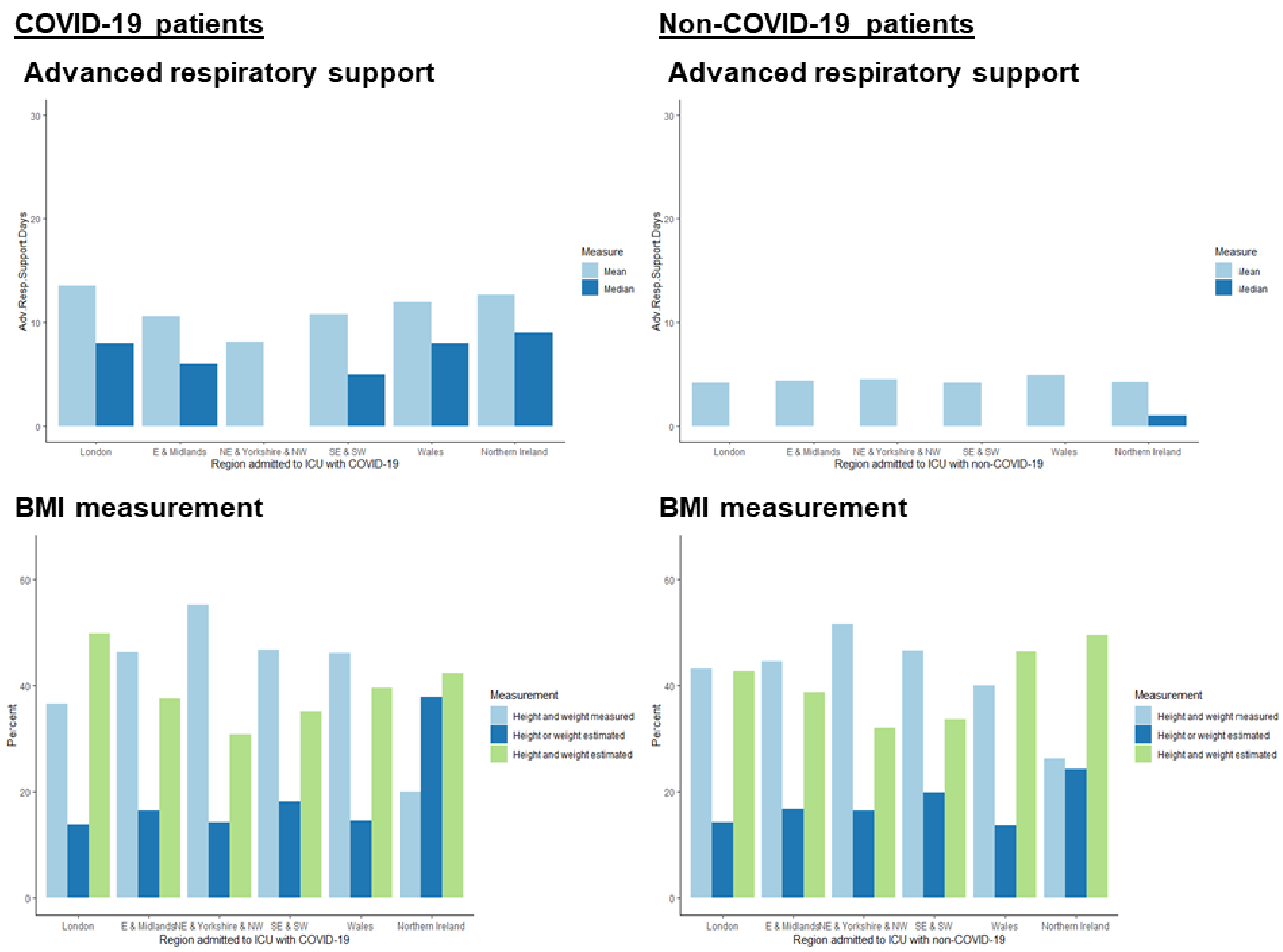


**Note:** Absent bars for advanced respiratory support indicate values of 0. E = East. NE = North-East. NW = North-West. SE = South-East. SW = South-West

**Supplementary Table 1** Associations of confounding and selection factors with BMI and 30-day all-cause mortality among ICU patients with COVID-19 (5 Feb 2020 to 1 Aug 2021) and non-COVID-19 respiratory conditions (1 Feb 2018 to 31 Aug 2019)

| | **COVID-19 patients** N=33,810 | **Non-COVID-19 patients** N=24,879 | |
|---|---|---|---|
| | **Mean difference (95% CI) in BMI ($kg/m^2$), adjusted for age and sex** | | **P-het. with patient group** |
| ***Socio-demographics*** | | | |
| Black vs. white ethnic group | -1.34 (-1.65, -1.03) | 0.03 (-0.54, 0.59) | <0.0001 |
| Asian vs. white ethnic group | -3.06 (-3.27, -2.85) | -1.35 (-1.79, -0.91) | <0.0001 |
| Mixed/other vs. white ethnic group | -2.22 (-2.52, -1.93) | -1.11 (-1.69, -0.53) | 0.001 |
| Deprivation (per quintile higher) | 0.15 (0.10, 0.21) | 0.18 (0.11, 0.24) | 0.62 |
| ***Prior or current comorbidities*** | | | |
| Any past severe illness | -1.22 (-1.48, -0.96) | -0.73 (-0.95, -0.51) | 0.01 |
| Some or total dependency pre-admission | 0.72 (0.48, 0.96) | 0.44 (0.25, 0.63) | 0.08 |
| Severe CVD | -0.46 (-1.43, 0.52) | 0.80 (0.14, 1.47) | 0.04 |
| End-stage renal disease | -1.99 (-2.57, -1.41) | -0.73 (-1.37, -0.09) | 0.004 |
| Severe respiratory disease | 1.67 (0.90, 2.43) | 0.51 (0.12, 0.90) | 0.01 |
| Liver disease | -3.02 (-4.00, -2.04) | -1.96 (-2.60, -1.32) | 0.08 |
| Metastatic disease | -2.39 (-3.34, -1.43) | -1.38 (-1.93, -0.83) | 0.07 |
| Haematological disease | -2.96 (-3.52, -2.40) | -1.17 (-1.62, -0.73) | <0.0001 |
| Immunocompromised | -2.00 (-2.40, -1.60) | -1.16 (-1.46, -0.85) | 0.001 |
| APACHE II acute severity score (per unit) | -0.08 (-0.10, -0.06) | -0.08 (-0.10, -0.07) | 0.84 |
| ICNARC acute severity score (per unit) | -0.002 (-0.01, 0.01) | -0.03 (-0.05, -0.02) | <0.0001 |
| PaO2/FiO2 ratio (per unit) | -0.15 (-0.15, -0.14) | -0.01 (-0.02, -0.00002) | <0.0001 |
| Adv. respiratory support days (per day) | 0.004 (-0.001, 0.01) | 0.02 (0.01, 0.02) | 0.04 |
| | **HR (95% CI) for mortality, adjusted for age and sex** | | **P-het. with patient group** |
| ***Socio-demographics*** | | | |
| Black vs. white ethnic group | 1.10 (1.02, 1.18) | 0.67 (0.55, 0.81) | <0.0001 |
| Asian vs. white ethnic group | 1.35 (1.29, 1.42) | 1.05 (0.93, 1.19) | 0.0001 |
| Mixed/other vs. white ethnic group | 0.92 (0.86, 0.99) | 0.75 (0.62, 0.91) | 0.05 |
| Deprivation (per quintile higher) | 1.04 (1.03, 1.06) | 1.00 (0.98, 1.02) | <0.0001 |
| ***Prior or current comorbidities*** | | | |
| Any past severe illness | 1.45 (1.37, 1.52) | 1.77 (1.67, 1.86) | <0.0001 |
| Some or total dependency pre-admission | 1.30 (1.24, 1.36) | 1.40 (1.33, 1.47) | 0.04 |
| Severe CVD | 1.37 (1.15, 1.63) | 1.58 (1.37, 1.83) | 0.21 |
| End-stage renal disease | 1.37 (1.22, 1.55) | 0.80 (0.66, 0.98) | <0.0001 |
| Severe respiratory disease | 1.42 (1.23, 1.63) | 1.50 (1.37, 1.65) | 0.49 |
| Liver disease | 2.33 (1.95, 2.78) | 2.66 (2.32, 3.04) | 0.25 |
| Metastatic disease | 1.53 (1.28, 1.82) | 1.91 (1.70, 2.14) | 0.04 |
| Haematological disease | 1.81 (1.64, 2.01) | 2.02 (1.84, 2.22) | 0.12 |
| Immunocompromised | 1.65 (1.53, 1.78) | 1.70 (1.58, 1.83) | 0.59 |
| APACHE II acute severity score (per unit) | 1.08 (1.08, 1.09) | 1.12 (1.11, 1.12) | <0.0001 |
| ICNARC acute severity score (per unit) | 1.07 (1.07, 1.07) | 1.10 (1.09, 1.10) | <0.0001 |
| PaO2/FiO2 ratio (per unit) | 0.95 (0.95, 0.96) | 0.95 (0.94, 0.95) | <0.0001 |
| Adv. respiratory support days (per day) | 0.99 (0.99, 0.99) | 1.00 (1.00, 1.00) | <0.0001 |

**Supplementary Table 2** Associations of confounding/selection factors with BMI among ICU patients with COVID-19, by admission date

| | **Feb-April 2020** N=8,054 | **May-July 2020** N=1,470 | **Aug-Oct 2020** N=2,878 | **Nov-Jan 2021** N=15,418 | **Feb-April 2021** N=4,061 | **May-Aug 2021** N=1,929 | |
|---|---|---|---|---|---|---|---|
| | **Mean difference (95% CI) in BMI (kg/m$^2$) among COVID-19 patients, adjusted for age and sex** | | | | | | **P-het. with date** |
| ***Socio-demographics*** | | | | | | | |
| Black vs. white ethnic group | -1.06 (-1.55, -0.57) | -0.86 (-2.58, 0.87) | -0.88 (-2.11, 0.36) | -1.46 (-1.96, -0.96) | -1.51 (-2.63, -0.39) | 0.08 (-1.12, 1.27) | 0.21 |
| Asian vs. white ethnic group | -2.98 (-3.39, -2.56) | -1.67 (-2.68, -0.65) | -2.27 (-2.95, -1.58) | -3.20 (-3.51, -2.88) | -3.77 (-4.44, -3.10) | -2.95 (-3.87. -2.03) | 0.003 |
| Mixed/other vs. white ethnic group | -1.89 (-2.41, -1.38) | -2.40 (-4.01, -0.78) | -3.02 (-4.20, -1.84) | -2.27 (-2.72, -1.82) | -2.15 (-3.12, -1.18) | -1.35 (-2.61, -0.08) | 0.41 |
| Deprivation (per quintile higher) | 0.01 (-0.09, 0.12) | 0.42 (0.15, 0.69) | 0.18 (-0.01, 0.36) | 0.13 (0.05, 0.21) | 0.25 (0.07, 0.42) | 0.21 (-0.04, 0.47) | 0.04 |
| ***Prior or current comorbidities*** | | | | | | | |
| Any past severe illness | -1.25 (-1.78, -0.73) | -1.98 (-3.07, -0.89) | -0.92 (-1.77, -0.08) | -1.15 (-1.54, -0.75) | -1.82 (-2.59, -1.05) | -0.41 (-1.51, 0.69) | 0.18 |
| Some or total dependency pre-admission | 0.88 (0.37, 1.38) | 0.16 (-0.82, 1.15) | 0.14 (-0.62, 0.90) | 0.78 (0.43, 1.14) | 0.52 (-0.19, 1.23) | 1.13 (-0.14, 2.40) | 0.47 |
| Severe CVD | 0.37 (-1.76, 2.50) | -2.04 (-5.44, 1.36) | 1.09 (-1.44, 3.62) | -0.85 (-2.34, 0.63) | -1.90 (-4.77, 0.98) | 1.56 (-3.92, 7.04) | 0.45 |
| End-stage renal disease | -1.51 (-2.65, -0.37) | 2.18 (-0.27, 4.63) | -1.52 (-3.56, 0.52) | -2.73 (-3.58, -1.87) | -2.74 (-4.48, -1.00) | -1.24 (-3.92, 1.43) | 0.01 |
| Severe respiratory disease | 1.15 (-0.47, 2.78) | -1.99 (-4.47, 0.49) | 1.87 (-0.31, 4.04) | 1.67 (0.44, 2.91) | 3.61 (1.35, 5.87) | 4.22 (1.10, 7.33) | 0.01 |
| Liver disease | 1.19 (-1.50, 3.88) | -5.57 (-8.86, -2.27) | -3.71 (-6.58, -0.85) | -2.91 (-4.36, -1.47) | -4.96 (-7.51, -2.42) | -4.27 (-8.74, 0.20) | 0.02 |
| Metastatic disease | -0.07 (-2.25, 2.11) | -4.47 (-7.86, -1.08) | -2.19 (-5.06, 0.68) | -2.61 (-3.98, -1.24) | -4.83 (-7.76, -1.91) | 0.65 (-4.26, 5.56) | 0.07 |
| Haematological disease | -2.93 (-4.03, -1.83) | -4.37 (-6.32, -2.42) | -2.05 (-3.99, -0.11) | -2.76 (-3.65, -1.88) | -3.49 (-5.04, -1.94) | -2.15 (-4.88, 0.58) | 0.55 |
| Immunocompromised | -1.77 (-2.56, -0.97) | -3.59 (-5.30, -1.88) | -2.15 (-3.34, -0.96) | -1.66 (-2.28, -1.04) | -2.92 (-4.10, -1.74) | -1.60 (-3.27, 0.07) | 0.16 |
| APACHE II acute severity score (per unit) | -0.01 (-0.04, 0.02) | -0.08 (-0.15, -0.01) | -0.05 (-0.10, 0.004) | -0.10 (-0.12, -0.08) | -0.11 (-0.16, -0.06) | -0.07 (-0.14, -0.003) | 0.0002 |
| ICNARC acute severity score (per unit) | 0.03 (0.01, 0.05) | -0.02 (-0.07, 0.03) | 0.04 (0.001, 0.07) | -0.01 (-0.03, 0.001) | -0.01 (-0.04, 0.02) | 0.03 (-0.02, 0.08) | 0.003 |
| PaO2/FiO2 ratio (per unit) | -0.11 (-0.12, -0.09) | -0.13 (-0.16, -0.10) | -0.16 (-0.19, -0.14) | -0.15 (-0.16, -0.13) | -0.16 (-0.18, -0.13) | -0.18 (-0.21, -0.14) | <0.0001 |
| Adv. respiratory support days (per day) | -0.003 (-0.01, 0.01) | 0.02 (0.001, 0.05) | 0.02 (0.004, 0.04) | 0.004 (-0.003, 0.01) | 0.01 (-0.0003, 0.03) | 0.04 (0.002, 0.08) | 0.01 |

**Supplementary Table 3** Associations of confounding/selection factors with BMI among ICU patients with non-COVID-19 respiratory conditions, by admission date

| | **Feb-April 2018** N=4,587 | **May-July 2018** N=3,266 | **Aug-Oct 2018** N=3,128 | **Nov-Jan 2019** N=5,236 | **Feb-April 2019** N=4,432 | **May-Aug 2019** N=4,230 | |
|---|---|---|---|---|---|---|---|
| | **Mean difference (95% CI) in BMI (kg/m$^2$) among non-COVID-19 patients, adjusted for age and sex** | | | | | | **P-het. with date** |
| ***Socio-demographics*** | | | | | | | |
| Black vs. white ethnic group | 1.02 (-0.21, 2.24) | -0.59 (-2.06, 0.88) | 0.22 (-1.28, 1.72) | -1.89 (-3.21, -0.57) | 1.52 (0.10, 2.94) | 0.06 (-1.30, 1.41) | 0.01 |
| Asian vs. white ethnic group | -1.61 (-2.56, -0.65) | -1.85 (-3.09, -0.60) | -1.14 (-2.35, 0.06) | -1.21 (-2.21, -0.22) | -1.54 (-2.61, -0.47) | -0.94 (-1.97, 0.10) | 0.87 |
| Mixed/other vs. white ethnic group | -2.80 (-4.14, -1.47) | -0.59 (-2.19, 1.01) | -1.49 (-3.00, 0.01) | 0.39 (-0.87, 1.66) | -1.01 (-2.39, 0.36) | -1.69 (-3.26, -0.12) | 0.03 |
| Deprivation (per quintile higher) | 0.22 (0.08, 0.37) | 0.17 (-0.01, 0.34) | 0.12 (-0.06, 0.30) | 0.09 (-0.07, 0.24) | 0.24 (0.08, 0.40) | 0.19 (0.02, 0.36) | 0.72 |
| ***Prior or current comorbidities*** | | | | | | | |
| Any past severe illness | -0.36 (-0.86, 0.15) | -0.73 (-1.31, -0.16) | -1.29 (-1.88, -0.70) | -0.97 (-1.48, -0.45) | -0.42 (-0.96, 0.12) | -0.67 (-1.22, -0.12) | 0.20 |
| Some or total dependency pre-admission | 0.38 (-0.05, 0.82) | 0.57 (0.06, 1.09) | 0.19 (-0.33, 0.72) | 0.16 (-0.29, 0.60) | 0.62 (0.15, 1.08) | 0.75 (0.27, 1.24) | 0.41 |
| Severe CVD | 1.91 (0.41, 3.40) | 0.45 (-1.20, 2.10) | 0.09 (-1.79, 1.96) | 0.94 (-0.64, 2.52) | 0.73 (-0.82, 2.29) | 0.33 (-1.30, 1.97) | 0.70 |
| End-stage renal disease | -1.18 (-2.55, 0.19) | 0.26 (-1.45, 1.97) | -0.54 (-2.47, 1.40) | -1.05 (-2.56, 0.47) | -1.03 (-2.54, 0.48) | -0.50 (-2.01, 1.00) | 0.84 |
| Severe respiratory disease | 0.84 (-0.04, 1.72) | 0.23 (-0.77, 1.23) | 0.29 (-0.75, 1.33) | -0.06 (-0.94, 0.81) | 1.19 (0.22, 2.16) | 0.71 (-0.27, 1.68) | 0.45 |
| Liver disease | -1.38 (-2.93, 0.16) | -1.16 (-2.72, 0.41) | -3.04 (-4.64, -1.45) | -1.60 (-3.20, 0.002) | -1.75 (-3.23, -0.26) | -2.71 (-4.30, -1.12) | 0.50 |
| Metastatic disease | -0.82 (-2.21, 0.57) | -1.32 (-2.71, 0.07) | -1.60 (-2.92, -0.29) | -1.90 (-3.27, -0.53) | -0.76 (-2.06, 0.54) | -1.68 (-2.98, -0.38) | 0.80 |
| Haematological disease | -0.69 (-1.73, 0.36) | -1.61 (-2.72, -0.50) | -1.04 (-2.25, 0.16) | -0.99 (-2.05, 0.07) | -1.17 (-2.22, -0.12) | -1.51 (-2.57, -0.45) | 0.86 |
| Immunocompromised | -0.65 (-1.36, 0.06) | -1.11 (-1.89, -0.33) | -1.73 (-2.53, -0.94) | -1.40 (-2.11, -0.69) | -1.05 (-1.79, -0.31) | -1.05 (-1.80, -0.30) | 0.51 |
| APACHE II acute severity score (per unit) | -0.08 (-0.12, -0.05) | -0.06 (-0.10, -0.02) | -0.11 (-0.15, -0.07) | -0.06 (-0.09, -0.02) | -0.08 (-0.11, -0.04) | -0.10 (-0.14, -0.06) | 0.30 |
| ICNARC acute severity score (per unit) | -0.05 (-0.07, -0.02) | -0.04 (-0.06, -0.01) | -0.04 (-0.06, -0.01) | 0.001 (-0.02, 0.03) | -0.03 (-0.06, -0.01) | -0.06 (-0.09, -0.03) | 0.02 |
| PaO2/FiO2 ratio (per unit) | -0.01 (-0.03, 0.01) | 0.01 (-0.02, 0.03) | 0.002 (-0.02, 0.03) | -0.03 (-0.05, -0.01) | -0.02 (-0.04, -0.003) | 0.02 (-0.001, 0.04) | 0.003 |
| Adv. respiratory support days (per day) | -0.001 (-0.02, 0.02) | 0.02 (-0.01, 0.04) | 0.03 (0.01, 0.06) | 0.02 (0.001, 0.04) | 0.03 (0.01, 0.05) | -0.003 (-0.03, 0.02) | 0.22 |

**Supplementary Table 4** Associations of confounding/selection factors with all-cause mortality among ICU patients with COVID-19, by admission date

| | **Feb-April 2020** N=8,054 | **May-July 2020** N=1,470 | **Aug-Oct 2020** N=2,878 | **Nov-Jan 2021** N=15,418 | **Feb-April 2021** N=4,061 | **May-Aug 2021** N=1,929 | |
|---|---|---|---|---|---|---|---|
| | **HR (95% CI) for mortality among COVID-19 patients, adjusted for age and sex** | | | | | | **P-het. with date** |
| ***Socio-demographics*** | | | | | | | |
| Black vs. white ethnic group | 1.29 (1.15, 1.44) | 1.04 (0.65, 1.65) | 0.82 (0.56, 1.21) | 0.98 (0.87, 1.10) | 0.95 (0.73, 1.23) | 0.86 (0.58, 1.27) | 0.01 |
| Asian vs. white ethnic group | 1.39 (1.27, 1.53) | 1.42 (1.12, 1.80) | 1.22 (1.04, 1.43) | 1.36 (1.27, 1.45) | 1.38 (1.20, 1.59) | 1.32 (1.01, 1.73) | 0.80 |
| Mixed/other vs. white ethnic group | 1.06 (0.93, 1.20) | 0.95 (0.60, 1.49) | 0.61 (0.42, 0.87) | 0.91 (0.82, 1.02) | 0.68 (0.52, 0.88) | 0.75 (0.47, 1.20) | 0.01 |
| Deprivation (per quintile higher) | 1.06 (1.04, 1.09) | 1.05 (0.98, 1.12) | 1.04 (0.99, 1.08) | 1.06 (1.04, 1.08) | 1.02 (0.98, 1.06) | 1.02 (0.94, 1.10) | 0.43 |
| ***Prior or current comorbidities*** | | | | | | | |
| Any past severe illness | 1.26 (1.13, 1.40) | 1.71 (1.37, 2.14) | 1.70 (1.44, 2.01) | 1.48 (1.38, 1.60) | 1.52 (1.32, 1.75) | 1.74 (1.35, 2.24) | 0.01 |
| Some or total dependency pre-admission | 1.27 (1.14, 1.41) | 1.19 (0.95, 1.49) | 1.48 (1.27, 1.74) | 1.32 (1.23, 1.41) | 1.29 (1.12, 1.48) | 1.95 (1.47, 2.60) | 0.05 |
| Severe CVD | 0.99 (0.66, 1.48) | 1.58 (0.87, 2.88) | 1.31 (0.82, 2.09) | 1.52 (1.16, 1.98) | 1.73 (1.07, 2.80) | 1.35 (0.34, 5.40) | 0.53 |
| End-stage renal disease | 1.24 (0.97, 1.59) | 0.75 (0.40, 1.41) | 1.81 (1.19, 2.76) | 1.56 (1.31, 1.84) | 1.21 (0.84, 1.74) | 1.54 (0.79, 2.98) | 0.15 |
| Severe respiratory disease | 1.44 (1.09, 1.91) | 1.49 (0.90, 2.45) | 2.04 (1.37, 3.04) | 1.52 (1.22, 1.90) | 1.18 (0.77, 1.80) | 0.79 (0.35, 1.77) | 0.30 |
| Liver disease | 1.68 (0.98, 2.90) | 2.39 (1.28, 4.49) | 2.27 (1.34, 3.85) | 2.19 (1.68, 2.85) | 3.06 (1.98, 4.71) | 10.45 (6.07, 21.57) | 0.0001 |
| Metastatic disease | 0.97 (0.60, 1.59) | 1.30 (0.62, 2.75) | 2.56 (1.59, 4.14) | 1.69 (1.32, 2.16) | 1.50 (0.92, 2.46) | 2.01 (0.83, 4.88) | 0.14 |
| Haematological disease | 1.35 (1.08, 1.67) | 2.34 (1.64, 3.33) | 1.51 (1.03, 2.21) | 2.02 (1.73, 2.36) | 2.20 (1.71, 2.85) | 2.23 (1.36, 3.66) | 0.01 |
| Immunocompromised | 1.36 (1.15, 1.60) | 2.14 (1.53, 2.98) | 1.89 (1.51, 2.36) | 1.67 (1.49, 1.88) | 1.69 (1.36, 2.10) | 2.68 (1.91, 3.77) | 0.01 |
| APACHE II acute severity score (per unit) | 1.07 (1.07, 1.08) | 1.10 (1.08, 1.12) | 1.09 (1.08, 1.10) | 1.08 (1.08, 1.09) | 1.09 (1.08, 1.10) | 1.13 (1.11, 1.15) | <0.0001 |
| ICNARC acute severity score (per unit) | 1.06 (1.06, 1.07) | 1.08 (1.07, 1.09) | 1.08 (1.07, 1.09) | 1.07 (1.06, 1.07) | 1.07 (1.06, 1.07) | 1.08 (1.07, 1.10) | <0.0001 |
| PaO2/FiO2 ratio (per unit) | 0.95 (0.95, 0.96) | 0.95 (0.94, 0.96) | 0.96 (0.95, 0.97) | 0.95 (0.95, 0.95) | 0.95 (0.94, 0.96) | 0.93 (0.91, 0.95) | 0.11 |
| Adv. respiratory support days (per day) | 0.97 (0.97, 0.98) | 0.99 (0.98, 1.00) | 1.00 (0.99, 1.00) | 0.99 (0.99, 0.99) | 1.00 (0.99, 1.00) | 1.02 (1.01, 1.03) | <0.0001 |

**Supplementary Table 5** Associations of confounding/selection factors with all-cause mortality among ICU patients with non-COVID-19 respiratory conditions, by admission date

| | **Feb-April 2018** N=4,587 | **May-July 2018** N=3,266 | **Aug-Oct 2018** N=3,128 | **Nov-Jan 2019** N=5,236 | **Feb-April 2019** N=4,432 | **May-Aug 2019** N=4,230 | |
|---|---|---|---|---|---|---|---|
| | **HR (95% CI) for mortality among non-COVID-19 patients, adjusted for age and sex** | | | | | | **P-het. with date** |
| ***Socio-demographics*** | | | | | | | |
| Black vs. white ethnic group | 0.70 (0.45, 1.07) | 0.48 (0.26, 0.90) | 0.61 (0.34, 1.11) | 0.71 (0.45, 1.13) | 0.71 (0.44, 1.15) | 0.72 (0.47, 1.12) | 0.92 |
| Asian vs. white ethnic group | 1.04 (0.78, 1.37) | 1.24 (0.89, 1.72) | 1.15 (0.83, 1.61) | 1.01 (0.77, 1.32) | 0.91 (0.67, 1.24) | 1.06 (0.80, 1.39) | 0.82 |
| Mixed/other vs. white ethnic group | 0.91 (0.59, 1.40) | 0.75 (0.44, 1.28) | 1.04 (0.67, 1.61) | 0.67 (0.44, 1.03) | 0.57 (0.35, 0.94) | 0.67 (0.39, 1.13) | 0.48 |
| Deprivation (per quintile higher) | 0.99 (0.95, 1.03) | 1.04 (0.99, 1.09) | 0.98 (0.93, 1.03) | 1.02 (0.98, 1.06) | 0.99 (0.95, 1.03) | 0.98 (0.93, 1.02) | 0.33 |
| ***Prior or current comorbidities*** | | | | | | | |
| Any past severe illness | 1.55 (1.36, 1.76) | 1.70 (1.47, 1.97) | 2.00 (1.72, 2.32) | 1.71 (1.51, 1.93) | 1.88 (1.65, 2.14) | 1.87 (1.64, 2.13) | 0.13 |
| Some or total dependency pre-admission | 1.38 (1.23, 1.56) | 1.45 (1.26, 1.66) | 1.31 (1.13, 1.52) | 1.43 (1.27, 1.60) | 1.51 (1.34, 1.70) | 1.32 (1.16, 1.49) | 0.63 |
| Severe CVD | 1.00 (0.67, 1.48) | 1.41 (0.97, 2.06) | 1.18 (0.74, 1.89) | 2.13 (1.57, 2.89) | 2.68 (1.99, 3.60) | 1.30 (0.89, 1.89) | 0.0003 |
| End-stage renal disease | 0.74 (0.48, 1.16) | 0.74 (0.43, 1.29) | 0.99 (0.56, 1.76) | 0.90 (0.58, 1.38) | 0.68 (0.41, 1.11) | 0.84 (0.55, 1.29) | 0.91 |
| Severe respiratory disease | 1.34 (1.06, 1.68) | 1.54 (1.21, 1.98) | 1.61 (1.24, 2.10) | 1.37 (1.11, 1.69) | 1.62 (1.30, 2.03) | 1.64 (1.31, 2.04) | 0.68 |
| Liver disease | 2.63 (1.88, 3.67) | 2.60 (1.85, 3.64) | 2.96 (2.11, 4.15) | 2.02 (1.42, 2.88) | 2.45 (1.79, 3.35) | 3.46 (2.56, 4.67) | 0.31 |
| Metastatic disease | 1.99 (1.48, 2.68) | 1.59 (1.16, 2.18) | 2.26 (1.71, 2.98) | 2.11 (1.62, 2.76) | 1.77 (1.33, 2.34) | 1.85 (1.43, 2.41) | 0.60 |
| Haematological disease | 1.65 (1.29, 2.11) | 2.03 (1.60, 2.57) | 1.94 (1.48, 2.54) | 1.82 (1.46, 2.28) | 2.26 (1.82, 2.81) | 2.42 (1.98, 2.97) | 0.18 |
| Immunocompromised | 1.57 (1.32, 1.87) | 1.76 (1.47, 2.11) | 1.90 (1.57, 2.30) | 1.75 (1.49, 2.05) | 1.50 (1.26, 1.79) | 1.79 (1.52, 2.12) | 0.47 |
| APACHE II acute severity score (per unit) | 1.11 (1.10, 1.12) | 1.12 (1.11, 1.13) | 1.12 (1.11, 1.13) | 1.12 (1.11, 1.13) | 1.12 (1.11, 1.13) | 1.12 (1.11, 1.13) | 0.67 |
| ICNARC acute severity score (per unit) | 1.09 (1.08, 1.10) | 1.09 (1.09, 1.10) | 1.10 (1.09, 1.11) | 1.10 (1.10, 1.11) | 1.09 (1.08, 1.10) | 1.09 (1.09, 1.10) | 0.03 |
| PaO2/FiO2 ratio (per unit) | 0.94 (0.94, 0.95) | 0.95 (0.94, 0.96) | 0.94 (0.93, 0.95) | 0.94 (0.94, 0.95) | 0.95 (0.94, 0.96) | 0.95 (0.94, 0.95) | 0.23 |
| Adv. respiratory support days (per day) | 0.99 (0.99, 1.00) | 1.00 (0.99, 1.00) | 1.00 (0.99, 1.01) | 1.00 (0.99, 1.00) | 1.00 (0.99, 1.01) | 1.00 (1.00, 1.01) | 0.61 |

**Supplementary Table 6** Associations of confounding/selection factors with BMI among ICU patients with COVID-19 (5 Feb 2020 to 1 Aug 2021), by geographical region

| | **London** N=7,859 | **E England & Midlands** N=8,328 | **NE & NW England, & Yorkshire** N=10,372 | **SE & SW England** N=5,332 | **Wales** N=1,299 | **Northern Ireland** N=620 | |
|---|---|---|---|---|---|---|---|
| | **Mean difference (95% CI) in BMI (kg/m$^2$) among COVID-19 patients, adjusted for age and sex** | | | | | | **P-het. with region** |
| ***Socio-demographics*** | | | | | | | |
| Black vs. white ethnic group | -0.04 (-0.46, 0.38) | -0.94 (-1.66, -0.23) | -2.09 (-2.98, -1.20) | -1.67 (-2.77, -0.56) | -1.69 (-5.97, 2.59) | -4.77 (-11.82, 2.28) | 0.0003 |
| Asian vs. white ethnic group | -2.44 (-2.82, -2.07) | -2.67 (-3.09, -2.26) | -2.76 (-3.19, -2.32) | -3.74 (-4.36, -3.13) | -2.16 (-4.01, -0.31) | -8.19 (-13.47, -2.91) | 0.003 |
| Mixed/other vs. white ethnic group | -1.30 (-1.73, -0.86) | -1.94 (-2.59, -1.28) | -2.48 (-3.24, -1.72) | -1.92 (-2.81, -1.02) | -1.57 (-4.19, 1.05) | 0.08 (-3.34, 3.51) | 0.10 |
| Deprivation (per quintile higher) | 0.20 (0.09, 0.32) | 0.09 (-0.02, 0.20) | 0.17 (0.07, 0.28) | 0.21 (0.07, 0.35) | -0.06 (-0.34, 0.23) | 0.32 (-0.14, 0.77) | 0.37 |
| ***Prior or current comorbidities*** | | | | | | | |
| Any past severe illness | -1.22 (-1.70, -0.74) | -0.96 (-1.48, -0.44) | -1.24 (-1.74, -0.74) | -1.43 (-2.12, -0.74) | -1.41 (-2.83, 0.01) | -0.13 (-2.48, 2.22) | 0.80 |
| Some or total dependency pre-admission | 0.16 (-0.30, 0.61) | 0.98 (0.48, 1.48) | 0.62 (0.17, 1.07) | 1.18 (0.58, 1.79) | 0.81 (-0.35, 1.98) | 3.90 (1.55, 6.26) | 0.004 |
| Severe CVD | -0.19 (-2.14, 1.77) | -0.47 (-2.33, 1.39) | -1.09 (-2.69, 0.50) | 0.77 (-2.93, 4.48) | -1.83 (-6.78, 3.11) | 11.87 (0.75, 23.00) | 0.21 |
| End-stage renal disease | -1.37 (-2.23, -0.50) | -1.52 (-2.83, -0.22) | -2.59 (-3.82, -1.36) | -1.55 (-3.13, 0.02) | -0.70 (-4.11, 2.71) | -5.29 (-11.74, 1.17) | 0.47 |
| Severe respiratory disease | 3.52 (1.90, 5.13) | 2.52 (0.99, 4.05) | 0.06 (-1.18, 1.31) | 1.92 (-1.14, 4.98) | 1.74 (-1.19, 4.67) | 0.35 (-6.12, 6.83) | 0.03 |
| Liver disease | -3.33 (-4.94, -1.73) | -2.60 (-4.77, -0.42) | -2.08 (-3.92, -0.25) | -4.00 (-6.82, -1.18) | 0.06 (-5.19, 5.30) | -10.51 (-19.59, -1.43) | 0.28 |
| Metastatic disease | -1.40 (-3.02, 0.21) | -2.91 (-4.87, -0.95) | -2.24 (-4.07, -0.40) | -2.76 (-5.22, -0.29) | -4.01 (-10.64, 2.61) | -6.10 (-21.87, 9.67) | 0.84 |
| Haematological disease | -2.55 (-3.50, -1.60) | -3.53 (-4.67, -2.40) | -2.74 (-3.91, -1.56) | -2.57 (-4.01, -1.13) | -2.36 (-5.46, 0.75) | -2.59 (-7.19, 2.01) | 0.85 |
| Immunocompromised | -1.76 (-2.49, -1.04) | -2.44 (-3.31, -1.58) | -1.78 (-2.55, -1.01) | -1.91 (-2.86, -0.96) | -3.46 (-5.54, -1.38) | 0.72 (-2.47, 3.90) | 0.22 |
| APACHE II acute severity score (per unit) | -0.04 (-0.07, -0.02) | -0.06 (-0.09, -0.03) | -0.10 (-0.13, -0.07) | -0.08 (-0.12, -0.03) | -0.09 (-0.17, -0.01) | -0.10 (-0.24, 0.05) | 0.13 |
| ICNARC acute severity score (per unit) | 0.004 (-0.01, 0.02) | 0.01 (-0.01, 0.03) | 0.02 (0.001, 0.04) | -0.01 (-0.03, 0.02) | 0.001 (-0.05, 0.05) | -0.05 (-0.14, 0.04) | 0.34 |
| PaO2/FiO2 ratio (per unit) | -0.11 (-0.12, -0.09) | -0.15 (-0.16, -0.13) | -0.15 (-0.17, -0.14) | -0.18 (-0.20, -0.15) | -0.12 (-0.16, -0.08) | -0.25 (-0.32, -0.18) | <0.0001 |
| Adv. respiratory support days (per day) | 0.002 (-0.01, 0.01) | 0.02 (0.01, 0.03) | 0.01 (0.003, 0.02) | 0.01 (-0.001, 0.02) | -0.01 (-0.04, 0.02) | -0.01 (-0.05, 0.03) | 0.12 |

E = East. NE = North-East. NW = North-West. SE = South-East. SW = South-West.

**Supplementary Table 7** Associations of confounding/selection factors with BMI among ICU patients with non-COVID-19 (1 Feb 2018 to 31 Aug 2019), by geographical region

| | **London** N=4,511 | **E England & Midlands** N=6,259 | **NE & NW England, & Yorkshire** N=7,261 | **SE & SW England** N=4,697 | **Wales** N=1,446 | **Northern Ireland** N=705 | |
|---|---|---|---|---|---|---|---|
| | **Mean difference (95% CI) in BMI (kg/m$^2$) among non-COVID-19 patients, adjusted for age and sex** | | | | | | **P-het. with region** |
| ***Socio-demographics*** | | | | | | | |
| Black vs. white ethnic group | 0.73 (0.04, 1.42) | 0.62 (-0.87, 2.11) | 2.08 (0.01, 4.14) | -0.17 (-2.22, 1.88) | -3.55 (-13.40, 6.29) | NE (low count) | 0.54 |
| Asian vs. white ethnic group | -0.72 (-1.37, -0.08) | -1.10 (-1.98, -0.22) | -1.53 (-2.59, -0.47) | -1.23 (-2.72, 0.26) | -2.38 (-6.43, 1.66) | -4.25 (-15.06, 6.57) | 0.77 |
| Mixed/other vs. white ethnic group | -0.32 (-1.08, 0.45) | -1.92 (-3.38, -0.47) | 0.16 (-1.59, 1.90) | -2.13 (-4.00, -0.26) | 2.05 (-1.99, 6.09) | 3.46 (-2.34, 9.26) | 0.06 |
| Deprivation (per quintile higher) | 0.33 (0.17, 0.49) | 0.18 (0.04, 0.31) | 0.06 (-0.06, 0.19) | 0.34 (0.19, 0.50) | 0.09 (-0.17, 0.36) | -0.32 (-0.74, 0.09) | 0.004 |
| ***Prior or current comorbidities*** | | | | | | | |
| Any past severe illness | -0.79 (-1.26, -0.33) | -0.55 (-1.00, -0.09) | -0.91 (-1.32, -0.50) | -0.69 (-1.24, -0.14) | 0.04 (-0.82, 0.91) | -0.19 (-1.75, 1.36) | 0.47 |
| Some or total dependency pre-admission | 0.02 (-0.41, 0.44) | 0.62 (0.23, 1.01) | 0.56 (0.19, 0.92) | 0.71 (0.24, 1.18) | 0.15 (-0.60, 0.91) | 0.05 (-1.19, 1.28) | 0.23 |
| Severe CVD | 0.56 (-0.96, 2.08) | 1.19 (-0.06, 2.45) | 0.52 (-0.60, 1.65) | 1.58 (-0.68, 3.85) | 0.44 (-1.69, 2.56) | -3.53 (-11.20, 4.13) | 0.77 |
| End-stage renal disease | 0.32 (-0.84, 1.48) | -0.60 (-1.88, 0.69) | -1.85 (-3.12, -0.58) | -0.82 (-2.47, 0.82) | 0.78 (-2.31, 3.87) | -0.59 (-6.06, 4.88) | 0.23 |
| Severe respiratory disease | -0.12 (-1.07, 0.83) | 1.59 (0.78, 2.41) | -0.03 (-0.66, 0.60) | 0.09 (-1.06, 1.24) | 1.48 (0.20, 2.77) | 1.23 (-1.88, 4.34) | 0.02 |
| Liver disease | -1.73 (-3.24, -0.22) | -2.20 (-3.49, -0.92) | -2.50 (-3.62, -1.38) | -0.76 (-2.40, 0.89) | -1.52 (-3.93, 0.89) | -4.68 (-8.94, -0.42) | 0.42 |
| Metastatic disease | -1.30 (-2.30, -0.29) | -1.81 (-2.95, -0.68) | -1.19 (-2.31, -0.07) | -0.79 (-2.11, 0.53) | -0.47 (-3.31, 2.36) | -1.20 (-4.75, 2.35) | 0.89 |
| Haematological disease | -0.81 (-1.66, 0.04) | -1.48 (-2.37, -0.58) | -1.16 (-2.02, -0.31) | -0.99 (-2.11, 0.13) | -1.01 (-3.01, 0.99) | -0.65 (-4.13, 2.83) | 0.94 |
| Immunocompromised | -1.29 (-1.92, -0.65) | -1.34 (-1.99, -0.69) | -1.27 (-1.85, -0.70) | -0.75 (-1.46, -0.04) | -0.29 (-1.49, 0.90) | -0.46 (-2.51, 1.59) | 0.53 |
| APACHE II acute severity score (per unit) | -0.09 (-0.13, -0.06) | -0.07 (-0.10, -0.04) | -0.09 (-0.12, -0.06) | -0.07 (-0.11, -0.04) | -0.06 (-0.12, -0.001) | -0.12 (-0.21, -0.02) | 0.77 |
| ICNARC acute severity score (per unit) | -0.05 (-0.07, -0.02) | -0.04 (-0.06, -0.02) | -0.03 (-0.05, -0.01) | -0.04 (-0.06, -0.01) | -0.04 (-0.08, 0.004) | -0.04 (-0.10, 0.03) | 0.95 |
| PaO2/FiO2 ratio (per unit) | -0.01 (-0.02, 0.01) | -0.01 (-0.02, 0.01) | -0.01 (-0.02, 0.01) | 0.003 (-0.02, 0.02) | -0.002 (-0.04, 0.04) | -0.03 (-0.08, 0.03) | 0.92 |
| Adv. respiratory support days (per day) | -0.01 (-0.03, 0.01) | 0.03 (0.01, 0.05) | 0.02 (-0.001, 0.03) | 0.02 (-0.01, 0.04) | 0.02 (-0.02, 0.06) | 0.02 (-0.04, 0.09) | 0.18 |

E = East. NE = North-East. NW = North-West. SE = South-East. SW = South-West.

**Supplementary Table 8** Associations of confounding/selection factors with all-cause mortality among ICU patients with COVID-19 (5 Feb 2020 to 1 Aug 2021), by geographical region

| | **London** N=7,859 | **E England & Midlands** N=8,328 | **NE & NW England, & Yorkshire** N=10,372 | **SE & SW England** N=5,332 | **Wales** N=1,299 | **Northern Ireland** N=620 | |
|---|---|---|---|---|---|---|---|
| | **HR (95% CI) for mortality among COVID-19 patients, adjusted for age and sex** | | | | | | **P-het. with region** |
| ***Socio-demographics*** | | | | | | | |
| Black vs. white ethnic group | 1.16 (1.04, 1.29) | 1.25 (1.07, 1.46) | 0.98 (0.78, 1.23) | 1.14 (0.85, 1.54) | 1.09 (0.45, 2.63) | 1.03 (0.14, 7.42) | 0.69 |
| Asian vs. white ethnic group | 1.47 (1.35, 1.61) | 1.32 (1.21, 1.44) | 1.31 (1.19, 1.44) | 1.44 (1.25, 1.66) | 1.10 (0.74, 1.66) | 1.24 (0.31, 5.06) | 0.34 |
| Mixed/other vs. white ethnic group | 0.95 (0.85, 1.07) | 0.93 (0.79, 1.09) | 0.88 (0.72, 1.08) | 1.15 (0.92, 1.44) | 1.16 (0.65, 2.07) | 1.29 (0.57, 2.95) | 0.51 |
| Deprivation (per quintile higher) | 1.04 (1.01, 1.07) | 1.04 (1.02, 1.07) | 1.04 (1.02, 1.07) | 1.04 (1.00, 1.07) | 1.07 (1.01, 1.14) | 0.99 (0.90, 1.09) | 0.84 |
| ***Prior or current comorbidities*** | | | | | | | |
| Any past severe illness | 1.18 (1.06, 1.31) | 1.60 (1.45, 1.75) | 1.50 (1.37, 1.65) | 1.53 (1.33, 1.77) | 1.36 (1.04, 1.78) | 1.98 (1.32, 2.98) | 0.0004 |
| Some or total dependency pre-admission | 1.34 (1.21, 1.47) | 1.11 (1.01, 1.22) | 1.40 (1.29, 1.53) | 1.44 (1.27, 1.64) | 1.33 (1.06, 1.66) | 1.24 (0.74, 2.05) | 0.01 |
| Severe CVD | 1.38 (0.96, 1.98) | 1.35 (0.99, 1.84) | 1.22 (0.89, 1.65) | 2.93 (1.57, 5.46) | 1.01 (0.38, 2.70) | 1.52 (0.21, 10.86) | 0.24 |
| End-stage renal disease | 1.18 (0.97, 1.44) | 1.61 (1.24, 2.08) | 1.45 (1.13, 1.85) | 1.37 (0.96, 1.94) | 1.87 (1.03, 3.40) | 1.98 (0.73, 5.37) | 0.38 |
| Severe respiratory disease | 1.78 (1.30, 2.45) | 1.25 (0.97, 1.60) | 1.28 (1.00, 1.63) | 2.95 (1.67, 5.20) | 1.11 (0.64, 1.94) | 3.19 (1.18, 8.63) | 0.02 |
| Liver disease | 2.34 (1.69, 3.24) | 2.25 (1.57, 3.23) | 1.97 (1.41, 2.77) | 3.37 (2.03, 5.61) | 3.64 (1.62, 8.20) | 2.03 (0.50, 8.30) | 0.52 |
| Metastatic disease | 1.07 (0.74, 1.54) | 1.56 (1.11, 2.19) | 1.91 (1.40, 2.62) | 2.28 (1.48, 3.50) | 0.68 (0.17, 2.75) | NE (low count) | 0.08 |
| Haematological disease | 1.58 (1.29, 1.92) | 1.88 (1.55, 2.29) | 1.91 (1.57, 2.33) | 1.94 (1.49, 2.52) | 2.39 (1.47, 3.87) | 1.50 (0.66, 3.38) | 0.55 |
| Immunocompromised | 1.39 (1.18, 1.63) | 1.75 (1.51, 2.04) | 1.91 (1.66, 2.19) | 1.63 (1.35, 1.98) | 1.45 (0.98, 2.13) | 1.75 (1.01, 3.03) | 0.08 |
| APACHE II acute severity score (per unit) | 1.08 (1.07, 1.08) | 1.09 (1.09, 1.10) | 1.08 (1.08, 1.09) | 1.09 (1.08, 1.10) | 1.07 (1.05, 1.08) | 1.11 (1.08, 1.15) | 0.0001 |
| ICNARC acute severity score (per unit) | 1.07 (1.06, 1.07) | 1.07 (1.06, 1.07) | 1.08 (1.07, 1.08) | 1.08 (1.07, 1.08) | 1.06 (1.05, 1.07) | 1.05 (1.03, 1.07) | 0.003 |
| PaO2/FiO2 ratio (per unit) | 0.96 (0.95, 0.96) | 0.96 (0.95, 0.96) | 0.95 (0.94, 0.95) | 0.95 (0.95, 0.96) | 0.95 (0.94, 0.97) | 0.93 (0.91, 0.95) | 0.004 |
| Adv. respiratory support days (per day) | 0.98 (0.98, 0.99) | 0.99 (0.99, 0.99) | 1.00 (0.99, 1.00) | 0.99 (0.99, 1.00) | 0.98 (0.98, 0.99) | 0.98 (0.97, 0.99) | <0.0001 |

E = East. NE = North-East. NW = North-West. SE = South-East. SW = South-West.

**Supplementary Table 9** Associations of confounding/selection factors with all-cause mortality among ICU patients with non-COVID-19 (1 Feb 2018 to 31 Aug 2019), by geographical region

| | **London** N=4,511 | **E England & Midlands** N=6,259 | **NE & NW England, & Yorkshire** N=7,261 | **SE & SW England** N=4,697 | **Wales** N=1,446 | **Northern Ireland** N=705 | |
|---|---|---|---|---|---|---|---|
| | **HR (95% CI) for mortality among non-COVID-19 patients, adjusted for age and sex** | | | | | | **P-het. with region** |
| ***Socio-demographics*** | | | | | | | |
| Black vs. white ethnic group | 0.78 (0.61, 1.01) | 0.68 (0.41, 1.13) | 0.72 (0.37, 1.38) | 0.67 (0.30, 1.50) | 2.23 (0.31, 15.93) | NE (low count) | 0.83 |
| Asian vs. white ethnic group | 1.12 (0.93, 1.36) | 1.11 (0.88, 1.39) | 1.10 (0.83, 1.47) | 1.25 (0.84, 1.86) | 1.28 (0.41, 4.00) | 2.27 (0.32, 16.32) | 0.98 |
| Mixed/other vs. white ethnic group | 0.90 (0.70, 1.16) | 0.78 (0.48, 1.28) | 0.88 (0.52, 1.49) | 0.64 (0.32, 1.28) | NE (low count) | 0.68 (0.09, 4.88) | 0.96 |
| Deprivation (per quintile higher) | 0.96 (0.92, 1.01) | 1.01 (0.98, 1.05) | 0.99 (0.96, 1.02) | 0.99 (0.94, 1.03) | 0.93 (0.87, 1.00) | 1.01 (0.89, 1.14) | 0.29 |
| ***Prior or current comorbidities*** | | | | | | | |
| Any past severe illness | 2.02 (1.78, 2.29) | 1.67 (1.50, 1.86) | 1.63 (1.48, 1.80) | 1.94 (1.70, 2.22) | 1.79 (1.45, 2.20) | 1.45 (0.98, 2.16) | 0.06 |
| Some or total dependency pre-admission | 1.57 (1.39, 1.78) | 1.29 (1.17, 1.43) | 1.41 (1.29, 1.55) | 1.49 (1.31, 1.69) | 1.27 (1.05, 1.55) | 1.12 (0.78, 1.61) | 0.11 |
| Severe CVD | 1.60 (1.11, 2.30) | 1.37 (1.04, 1.82) | 1.65 (1.30, 2.08) | 1.73 (1.07, 2.79) | 1.33 (0.81, 2.19) | 2.57 (0.63, 10.45) | 0.84 |
| End-stage renal disease | 0.86 (0.60, 1.25) | 0.88 (0.61, 1.27) | 0.90 (0.64, 1.28) | 0.33 (0.15, 0.74) | 0.51 (0.16, 1.60) | 6.49 (2.56, 16.48) | 0.0002 |
| Severe respiratory disease | 1.77 (1.38, 2.27) | 1.29 (1.06, 1.57) | 1.37 (1.18, 1.59) | 2.07 (1.61, 2.66) | 1.18 (0.85, 1.64) | 1.39 (0.65, 2.98) | 0.02 |
| Liver disease | 2.44 (1.72, 3.45) | 2.45 (1.87, 3.20) | 3.02 (2.42, 3.75) | 2.57 (1.80, 3.66) | 1.72 (0.97, 3.07) | 5.28 (2.45, 11.39) | 0.21 |
| Metastatic disease | 1.82 (1.44, 2.32) | 1.66 (1.30, 2.12) | 2.20 (1.79, 2.72) | 2.09 (1.59, 2.75) | 2.94 (1.76, 4.93) | 1.74 (0.77, 3.95) | 0.31 |
| Haematological disease | 2.32 (1.91, 2.81) | 2.07 (1.73, 2.49) | 1.79 (1.50, 2.14) | 2.08 (1.64, 2.64) | 2.51 (1.71, 3.69) | 1.28 (0.56, 2.91) | 0.30 |
| Immunocompromised | 1.94 (1.65, 2.28) | 1.72 (1.49, 1.99) | 1.43 (1.25, 1.63) | 2.01 (1.72, 2.36) | 1.91 (1.47, 2.48) | 0.94 (0.52, 1.71) | 0.003 |
| APACHE II acute severity score (per unit) | 1.13 (1.11, 1.14) | 1.11 (1.10, 1.12) | 1.12 (1.11, 1.13) | 1.12 (1.11, 1.13) | 1.10 (1.08, 1.12) | 1.17 (1.14, 1.20) | 0.001 |
| ICNARC acute severity score (per unit) | 1.10 (1.09, 1.11) | 1.09 (1.09, 1.10) | 1.10 (1.09, 1.10) | 1.10 (1.09, 1.11) | 1.08 (1.07, 1.09) | 1.12 (1.10, 1.14) | 0.01 |
| PaO2/FiO2 ratio (per unit) | 0.95 (0.95, 0.96) | 0.95 (0.95, 0.96) | 0.94 (0.94, 0.95) | 0.94 (0.93, 0.95) | 0.94 (0.92, 0.95) | 0.93 (0.91, 0.95) | 0.01 |
| Adv. respiratory support days (per day) | 1.00 (1.00, 1.00) | 1.00 (0.99, 1.00) | 0.99 (0.99, 1.00) | 1.01 (1.00, 1.01) | 0.99 (0.98, 1.00) | 1.01 (1.00, 1.03) | 0.002 |

E = East. NE = North-East. NW = North-West. SE = South-East. SW = South-West.
